# Mitigating the impact of motor impairment on self-administered digital tests in patients with neurological disorders

**DOI:** 10.1101/2025.03.14.25323903

**Authors:** Dragos-Cristian Gruia, Valentina Giunchiglia, Andra Braban, Niamh Parkinson, Soma Banerjee, Joseph Kwan, Peter J. Hellyer, Adam Hampshire, Fatemeh Geranmayeh

## Abstract

**Introduction:** Cognitive impairments are prevalent in many neurological disorders and remain underdiagnosed and poorly studied longitudinally. Unsupervised remote cognitive testing is an accessible, scalable, and cost-effective solution, however it often fails to separate cognitive deficits from commonly co-occurring motor impairments. To address this gap, we present a computational framework that isolates cognitive ability from motor impairment in self-administered digital tasks.

**Methods:** Stroke was chosen as a representative neurological disorder, as patients frequently experience both motor and cognitive impairments. Our validation analyses spanned 18 computerised tasks completed by 171 patients longitudinally, covering a broad spectrum of cognitive and motor domains. The computational model was applied on trial-level data to disentangle the contribution of motor and cognitive processes.

**Results:** In patients with motor hand impairment, standard accuracy performance metrics were confounded in 6 tasks (p<.05, FDR-corrected). In contrast, the Modelled Cognitive metrics obtained from the computational framework showed no significant effects of impaired hand (p>.05, FDR-corrected). Moreover, the Modelled Cognitive metrics correlated more strongly with clinical pen-and-paper scales (mean R^2^=0.64 vs. 0.43) and functional outcomes (mean R^2^=0.16 vs 0.09). Brain-behaviour associations were stronger when using the Modelled Cognitive metrics, and revealed intuitive multivariate relationships with individual tasks.

**Interpretation:** We present converging evidence for the improved clinical utility and validity of the Modelled Cognitive metrics within neurological conditions characterised by co-occurring motor and cognitive deficits. Addressing the confounding effect of motor impairments improves the reliability and biological validity of self-administered digital assessments, enhances accessibility, and supports early detection and intervention across neurological disorders.

**Funding:** This research is funded by the UK Medical Research Council (MR/T001402/1).

## Introduction

Remote digital cognitive assessments are poised to transform clinical research and healthcare. A growing array of digital technologies is being rolled out to support neurological care across the disease severity continuum, from presymptomatic stages to end-of-life care^1^. Such remote assessments have shown promise in enhancing the detection of neurological events, monitoring disease trajectory and progression, evaluating the effectiveness of clinical interventions, and stratifying patients into rehabilitation trials^2–4^. Moreover, the development and application of these technologies in routine care, and specifically applications for diagnosing cognitive impairment, have been highlighted as priority research areas by stakeholders^5,6^.

Self-administered digital cognitive assessments present several significant advantages over traditional pen-and-paper methods. Notably, they can be conducted unsupervised or semi-supervised, reducing the number of clinical visits required during patient’s journey, thereby, decreasing the burden on patients, healthcare providers and researchers^7,8^. Leveraging widely accessible smart devices or computers, these assessments eliminate the need for a clinician to be present, enabling cost-effective and scalable implementation. In contrast with most pen-and-paper assessments, digital tests generate detailed and standardised outcomes, facilitating harmonisation across different sites and reducing assessor bias.

Despite their advantages, these digital technologies come with notable challenges, largely pertaining to the unsupervised administration settings. First, is the inability to differentiate effects of cognitive and motor impairments commonly co-occurring in neurological conditions such as Parkinson’s Disease, Traumatic Brain Injury, Stroke, and Multiple Sclerosis^9–11^. Such differentiation is highly relevant for clinical interpretation and diagnosis as impaired motor skills can adversely affect metrics intended to measure cognitive performance. Current approaches have either overlooked this problem or tried to solve it by (i) excluding patients with motor impairments—who could benefit most from remote nature of the assessments—or (ii) excluding speed-based measures and focusing only on accuracy^12^. This latter approach discards large amounts of rich and potentially valuable data. Moreover, it is often unsuccessful when both the measures of accuracy and speed are inherently reliant on motor abilities, such as it is often the case when measuring processing speed and attention^10^.

An overlooked advantage of digital cognitive tasks over pen-and-paper tests, which can be exploited to overcome the above challenges, is their ability to record detailed trial-by-trial performance time-courses for each stimulus and response. This trial-by-trial performance variability can be modelled to derive ‘ability’ estimates with greater reliability and specificity than those obtained through simple averaging or summing of correct trials. One such computational framework, IDoCT (Iterative Decomposition of Cognitive Tasks), has recently been introduced in the context of neurologically healthy adults performing remote cognitive testing^13,14^. Briefly put, the IDoCT model examines trial-level variability in cognitive performance to generate a modelled ‘Cognitive Index’ that triangulates trial accuracy, reaction time, and trial difficulty, whilst mitigating confounding effects related to motoric issues, device variability and speed-accuracy trade-offs. This framework is designed to disentangle individuals’ cognitive abilities from the afore-mentioned confounding factors in a manner that is adaptive, robust, computationally efficient and compatible with known brain-behaviour associations^14^.

Whilst proof-of-concept was previously demonstrated in neurologically healthy cohorts, its clinical utility and validity in neurological populations with co-existing hand-motor and cognitive impairments remain unknown. Furthermore, the advantage and added clinical utility of IDoCT-derived cognitive measures, over traditional accuracy-based approaches, has not been explored. The current work aims to address this gap by applying the IDoCT model for the first time to a longitudinal cohort of 171 stroke survivors exhibiting a wide range of co-occurring cognitive and motor deficits.

First, the associations of the model-derived Cognitive Index and standard accuracy-based measures with demographic and clinical variables are compared separately to demonstrate intuitive relationships. Second, we investigate the task-specific effects of impaired hand motor function on cognitive performance across a diverse set of 18 self-administered cognitive tasks. We hypothesise that unlike conventional accuracy-based metrics, the modelled Cognitive Index will not be confounded by motor impairments. Third, we assess the clinical utility and validity of the modelled Cognitive Index against standard clinical scales (Montreal Cognitive Assessment - MoCA), functional outcomes following stroke (Instrumental Activities of Daily Living - IADL), and neuroimaging measures of cerebrovascular brain injury, including stroke lesion volume and white matter hyperintensity volume.

## Method

### Participants

#### Patient cohort

A total of 171 patients with radiologically confirmed stroke were consecutively recruited from Imperial College Healthcare NHS Trust. Out of these, 95 patients were assessed longitudinally after stroke, across the acute (within ∼1 week), sub-acute (∼3 months) and chronic (>6 months) recovery stages post-stroke (Table 1). Exclusion criteria included pre-stroke diagnosis of dementia, pure brain stem stroke, severe visuospatial problems interfering with cognitive testing, severe psychiatric co-morbidity, significant fatigue limiting engagement with cognitive testing. The digital task scores were related to clinical pen-and-paper cognitive screens (MoCA scores corrected for educational abilities, available for N=141), functional outcomes post-stroke (IADL, available for N=159) and magnetic resonance imaging (MRI) measures of cerebrovascular disease severity (stroke lesion volume and white matter hyperintensity volume, available in N=78).

**Table 1.** Demographic and clinical characteristics of the normative (N=6364) and stroke (N=171) cohorts.

|  | Neurologically<br>Healthy Cohort | Patients with stroke cohort |
| --- | --- | --- |
| **Number of participants:**<br>Whole group | N=6364 | N=171<br>Acute phase: N=132<br>Sub-acute phase: N=72<br>Chronic phase: N=131 |
| **Days since stroke:** (Median:IQR)<br>Acute<br>Subacute<br>Chronic phase |  | 4:2-7<br>94:88-104<br>371:217-445 |
| **Age in years:**<br>(Mean±SD; Range) | 60.8±10.1; 40-95 | 63.01±14.1; 24-97 |
| **\*Biological sex:**<br>(%Male:Female) | 42.7%:57.3% | 69.6%:30.4% |
| **\*Education %:** |  |  |
| Secondary school ≤16 years old (GCSE level) | 15.5% (N=984) | 8.2% (N=14) |
| Secondary school 18 years old (A-level) | 37.2% (N=2364) | 50.3% (N=86) |
| Bachelor's degree | 28.8% (N=1835) | 22.8% (N=39) |
| Postgraduate Degree | 18.5% (N=1181) | 18.7% (N=32) |
| **English as second language %** | 3.6% (N=231) | 34.5% (N=59) |
| **Stroke representation** |  |  |
| **% Stroke Aetiology:** |  |  |
| Ischaemic |  | 82.9% |
| Haemorrhagic |  | 17.1% |
| **% Hemisphere affected:** |  |  |
| Left |  | 53.7% |
| Right |  | 38.4% |
| Bilateral |  | 7.9% |
| **MOCA:** (Mean±SD; Range) |  |  |
| Acute |  | 20.96±6.09; 1-30 |
| Subacute |  | 22.70±6.15; 6-30 |
| Chronic phase |  | 24.19±5.25; 6-30 |
| **Stroke severity at baseline (NIHSS)** (Mean±SD; Range) |  | 4.93±4.66; 0-26 |
| Note. MOCA=Montreal Cognitive Assessment. NIHSS= National Institute of Health Stroke Scale, higher values relate to more severe stroke. “**” indicated that patients and controls differed significantly in their distributions. Total number of unique stroke survivors is indicated as 171. These patients had multiple assessments longitudinally. |  |  |

#### Normative cohort

Following participant exclusion and pre-processing steps, the final normative cohort used in this study contained 6364 healthy older adults described previously^15^.

All participants gave informed consent. The data was acquired as part of a longitudinal observational clinical study approved by UK’s Health Research Authority (Registered under NCT05885295; IRAS:299333; REC:21/SW/0124)^16^.

### Digital tasks

The data was obtained via the Imperial Comprehensive Cognitive Assessment in Cerebrovascular Disease (IC3), comprising 22 short tasks, specifically designed to cover a wide range of domains known to be affected after stroke; memory, language, executive function, attention, numeracy and praxis, as well as hand-motor dexterity^16–18^. We have previously demonstrated that IC3 is reliable, valid, and well-tolerated by patients in the acute and chronic stages after stroke^15^. The IC3 cognitive battery is deployable through a web-browser via a weblink in a device agnostic manner. The design and development of the tasks were performed in collaboration with Cognitron which also hosts the IC3 (https://www.cognitron.co.uk).

The healthy cohort and patients performed the tests in an unsupervised and semi-supervised manner respectively. Of the 22 tasks of IC3, 4 specifically assessed speech production abilities using recorded speech. Given our focus on differentiating the contribution of the hand-motor impairments from that of cognition, we excluded these 4 tasks for the purpose of this paper. All remaining 18 cognitive tasks were analysed. For the purpose of brevity, and to facilitate interpretations of the results, the main body of the manuscript focuses on results from four representative tasks (Figure 1) selected based on their varying levels of cognitive and motor demands (with the remaining 14 discussed as supplementary material). These four representative tasks include (1) the Motor Control task, characterised by high motor demand and low cognitive demand; (2) the Trail-Making task, characterised by high motor and high cognitive demand; (3) the Semantic Judgement task, characterised by low motor demand and high cognitive demand; and (4) the Simple Reaction Time (SRT) task, characterised by low motor demand and low cognitive demand. The details of the four tasks are shown in Figure 1. Further information on the remaining tasks, including their accuracy-based outcome measures has been previously described^15^.

**Figure 1.**
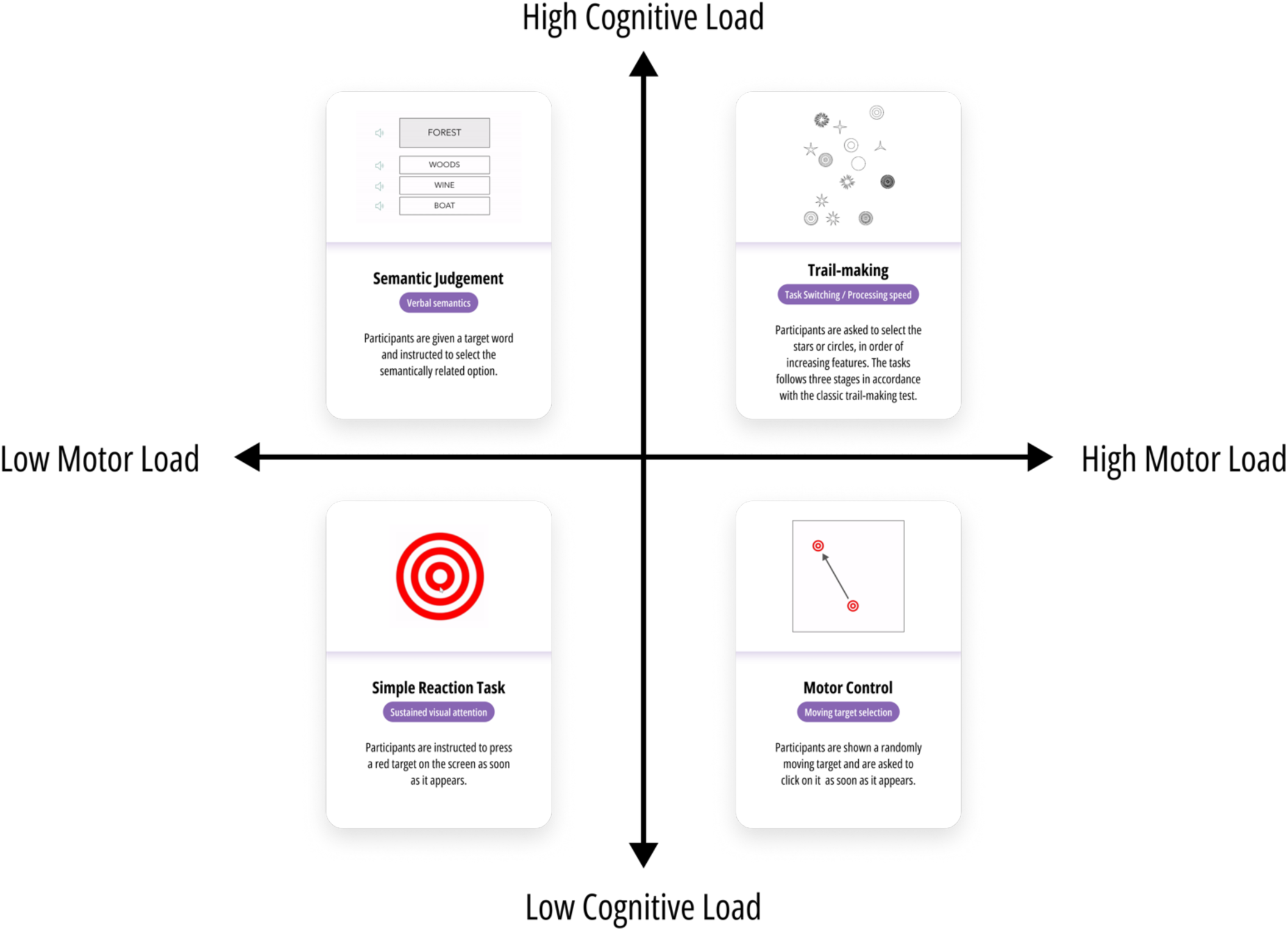
Four representative tasks selected from the self-administered IC3 cognitive battery, with opposing levels of motor and cognitive load. Each task is ∼3 minutes in duration. Highlighted text refers to the primary cognitive domain being assessed. The main outcome measure for each task is the total number of correct responses.

In addition to the above task-specific accuracy-based outcome measures, a summary performance metric for the entire cognitive battery was derived using a Bayesian principal component analysis (PCA)^19^. The analysis was conducted separately for data in the acute, sub-acute and chronic stages, as previous work has shown that the relationship between cognitive domains varies across the recovery phases^18,20^.

The advantage of using Bayesian PCA, over frequentist PCA was the ability to model missing data more confidently^19^. The algorithm employs an Expectation-Maximisation (EM) approach in conjunction with a Bayesian model to approximate the principal axes, which correspond to the eigenvectors of the covariance matrix in the PCA. The estimation process is iterative, and the algorithm terminates when the incremental improvement in precision becomes negligible at <10^-4^. The PCA-derived principal factor (Global factor G) explaining the largest proportion of variance was extracted separately for both the standard accuracy metrics (G_Standard Accuracy_) and the modelled Cognitive Indices (G_Modelled Cognitive Index_), representing overall performance on the battery. Both of these global cognitive measures (G_Standard Accuracy_ and G_Modelled Cognitive Index_) loaded positively on all tasks across all stages of recovery, explaining on average 59.17% (G_Standard Accuracy_: acute=60.6%, sub-acute=57.8%, chronic=59.1%) and 48.17% (G_Modelled Cognitive Index_: acute=44.5%, sub-acute=50.7%, chronic=49.3%) of the variance in global cognition respectively.

### Pre-processing steps for digital cognitive tasks

#### Normative cohort pre-processing

7,095 healthy older adults provided their consent and initiated the IC3 cognitive battery, with 5,639 participants (∼80%) successfully completing all 18 tasks. In order to ensure that the normative data was derived only from healthy participants who were fully engaged, several levels of data filtering was applied as discussed previously^15^. In brief, participants with neurological comorbidities or with family history of young-onset dementia were excluded (N=498).

Within participants, any individual task, or trials, where participants showed signs of non-engagement, (for example based on interactions outside the browser, and physiologically implausible reaction times), were additionally removed. In total, 10.1% of task-level data was excluded. The final cleaned normative sample contained 6364 individuals.

#### Patient cohort pre-processing

During the self-administration of the tasks, a trained researcher would flag any instances where any technical issues occurred (e.g., task did not load due to loss of internet connection) or the patient did not fully engage in the task (e.g., patient was interrupted part-way through the task by clinical staff). This information was used to exclude specific tasks from further analysis. In total, 4.15% of task-level data was removed.

### Computational modelling of cognitive data

The mathematical model involves several steps^13^. First, using the large age-matched normative cohort, the IDoCT model incorporates task-specific labels for each trial, and subsequently calculates a difficulty level for each trial relative to the labels provided. For instance, in the Semantic Judgement task, where each trial involves matching words of increasing semantic ambiguity, each target word is assigned a unique condition label for which a difficulty level is calculated (scaled 0-1).

Second, an iterative process is implemented based on the assumption that the difficulty of each trial (denoted as *D* in the original publication) can be determined from the group’s average performance (denoted as *P* in the original publication) on that trial^14^. Group performance, in turn, is calculated based on both reaction time (RT), accuracy of all participants and the difficulty of the trial. The iterative process continues until the trial difficulty and group performance estimates converge to stable values. Following this, a second iterative procedure is conducted, wherein the previously derived difficulty (*D)* values are used to estimate the answer time (*AT*). This second iteration assumes that RT consists of two components: the answer time (*AT*), representing the time required to complete the cognitive task, and the Response Delay Time (DT), which accounts for delays introduced by motor response, device latency, or other non-cognitive processes. Once *AT* is calculated, *DT* is obtained as the average difference between RT and *AT* across all trials. Simultaneously, the Cognitive Index (referred to as *AS* in the original publication) is computed as the cumulative measure of task-specific performance, focusing exclusively on the cognitive component of RT (i.e., *AT*). Additionally, a measure of scaled difficulty is derived, in which the initial trial difficulty *D* is adjusted according to the abilities of all the participants who completed those trials. This scaling is critical to mitigate biases that may arise when participants with higher cognitive abilities are disproportionately likely to progress to more challenging trials in case of cognitive tasks without random sampling of trials^13^. For transparency, the labels assigned to each trial for each task, along with the model’s estimated scaled difficulty for each label, are presented in Supplementary Figure 1. Overall, the assigned difficulty values were interpretable across all 18 tasks and demonstrated strong alignment with the intended task design and its associated cognitive demands. Task-level distributions for standard and modelled metrics can also be found in Supplementary Figure 2.

The modelled Cognitive Index captures aspects of trial accuracy, whilst integrating response speed and the difficulty of each trial. Thus, two participants with comparable raw accuracy may show a different modelled Cognitive Index if one makes slower responses and/or if the trials they completed are estimated to be more difficult. The modelled Response Delay Time captures individual differences in fundamental visuo-motor latencies (for example due to motor hand impairment, visual presentation on the device, delays caused by device latency or other non-cognitive processes), rather than task-related cognitive processes^13,14^.

### Mixed-effects regression analyses of the longitudinal sample

Separate linear mixed-effects regression models were employed to investigate the relationship between cognitive metrics (Modelled Cognitive Index and Standard Accuracy) and demographic and clinical variables of interest. Due to the longitudinal nature of the dataset, it was essential to account for repeated measurements from individual patients and to allow the cognitive recovery slopes to vary as a function of time. This approach was adopted because patients exhibit heterogeneous recovery trajectories following a stroke, and that this trajectory is often dependent on the initial stroke severity^18,20,21^. To examine the association between cognition and various demographic and clinical variables, we used the following regression model based on parameters described in Table 1:

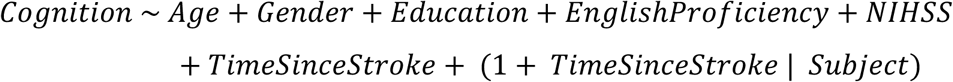

All dependent variables as well continuous covariates were standardised except for stroke severity (NIHSS) which was log transformed due its skewed distribution. A further mixed-effects analysis was conducted separately for each task using the same regression model described above, with hand motor impairment as an additional independent variable. This enabled assessment of the magnitude of the effect hand-motor impairment on the task performance whilst accounting for other confounding variables.

### Association between model derived cognitive metrics and measures of cerebrovascular disease brain injury

The global cognitive outcomes (G_Standard Accuracy_ and G_Modelled Cognitive Index_) were correlated against volumetric measures of brain tissue damage as defined by White Matter Hyperintensities (WMH) and Lesion volume on MRI brain scans derived from the subacute and chronic phase of stroke. Details of MRI data acquisition and calculations of the volumetric measures of WMH and stroke lesion are shown in Supplementary Methods.

We further confirmed meaningful relationships between cognitive performance and MRI measures of cerebrovascular disease severity using a multivariate Canonical Correlation Analysis (CCA). This method finds the strongest relationships between the two groups of variables (imaging and cognition), while Pearson correlation only identifies simple one-to-one relationships^22^.

## Results

### Participant cohorts

#### Participant characteristics

The data used for analysis consisted of 171 patients with stroke and 6364 neurologically healthy older adults. Of the 171 patients included, 95 were followed up longitudinally. Table 1 outlines the demographic and clinical characteristics of the two groups. Supplementary Figure 6 shows the distributions of the stroke lesions and white matter hyperintensities in the patient cohort.

### Effects of demographic and clinical factors on cognitive outcome measures

Separate mixed-effects linear regressions were performed to assess the interpretability of the novel modelled Cognitive Index in comparison to standard accuracy measures. The relationship between each cognitive measure and relevant demographic and clinical factors were estimated separately as standardised beta coefficients for each task (Figure 2). Overall, it was observed that the direction of effects remained consistent irrespective of the cognitive outcome measure examined (i.e., the modelled Cognitive Index or standard accuracy scores). In general, age and stroke severity at baseline were negatively associated with cognition, whilst increasing levels of education and time since stroke generally exhibited positive associations. These findings suggest that the modelled metrics are interpretable and align with standard accuracy measures.

**Figure 2.**
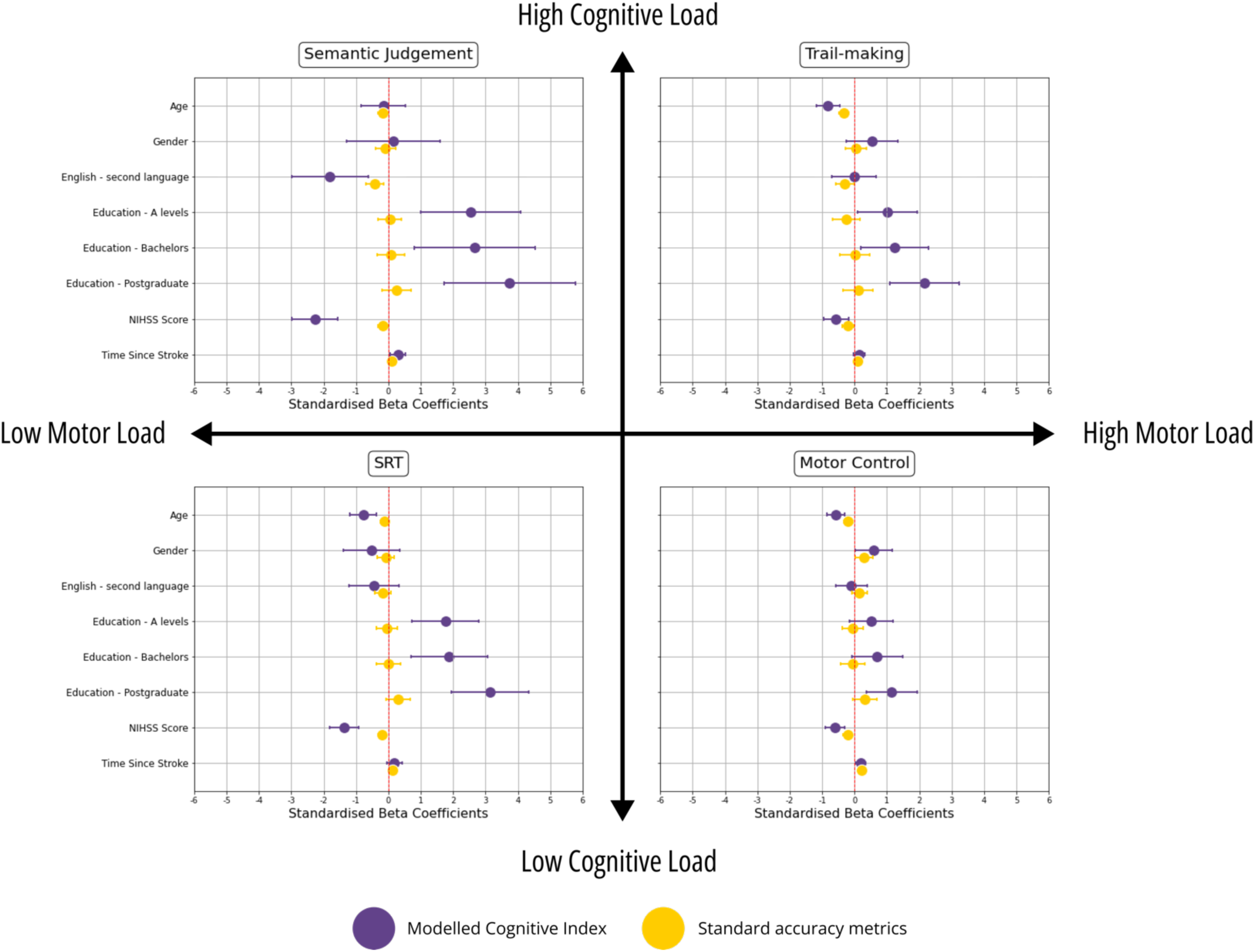
Relationship between cognitive scores and demographic and clinical variables in four representative tasks. Modelled Cognitive Indices and standard accuracy scores are displayed in purple and yellow respectively. The strength of the association is shown in standard deviation units, along with 95% confidence intervals. Asterisk denotes significantly stronger associations between the Modelled Cognitive Index and clinical variables compared to standard accuracy metrics. The reference value for education refers to patients with lowest education level (left education aged ≤16 years).

However, several notable differences were also observed. Specifically, in contrast to standard accuracy scores, the novel Cognitive Index demonstrated significantly stronger associations with several demographic and clinical factors such as education and stroke severity at baseline across a broad range of tasks, evident as increased separation of the confidence intervals shown in Figure 2. Overall, the modelled Cognitive Index was found to provide stronger associations with intuitive interpretations. A detailed breakdown of all 18 tasks is available in Supplementary Figure 3.

### The modelled Cognitive Index is independent of hand motor impairment

Hand motor impairment was defined as self-reported impairment in function of the hand used to make responses during the self-administration of the cognitive battery (N=47). This was verified against expected hand impairment based on stroke lesion location. The impact of using an impaired hand was assessed by including it as an additional predictor variable in the afore-mentioned mixed-effects regression models.

Standard accuracy metrics derived from tasks that had a high motor demand (trail making and motor control) were confounded by the presence of impaired hand function (P<.05, FDR-corrected, Figure 3). Overall, across 7 of the 18 self-administered tasks, participants performed significantly worse or responded more slowly when using their impaired hand, based on standard performance measures (Supplementary Figure 4).

**Figure 3.**
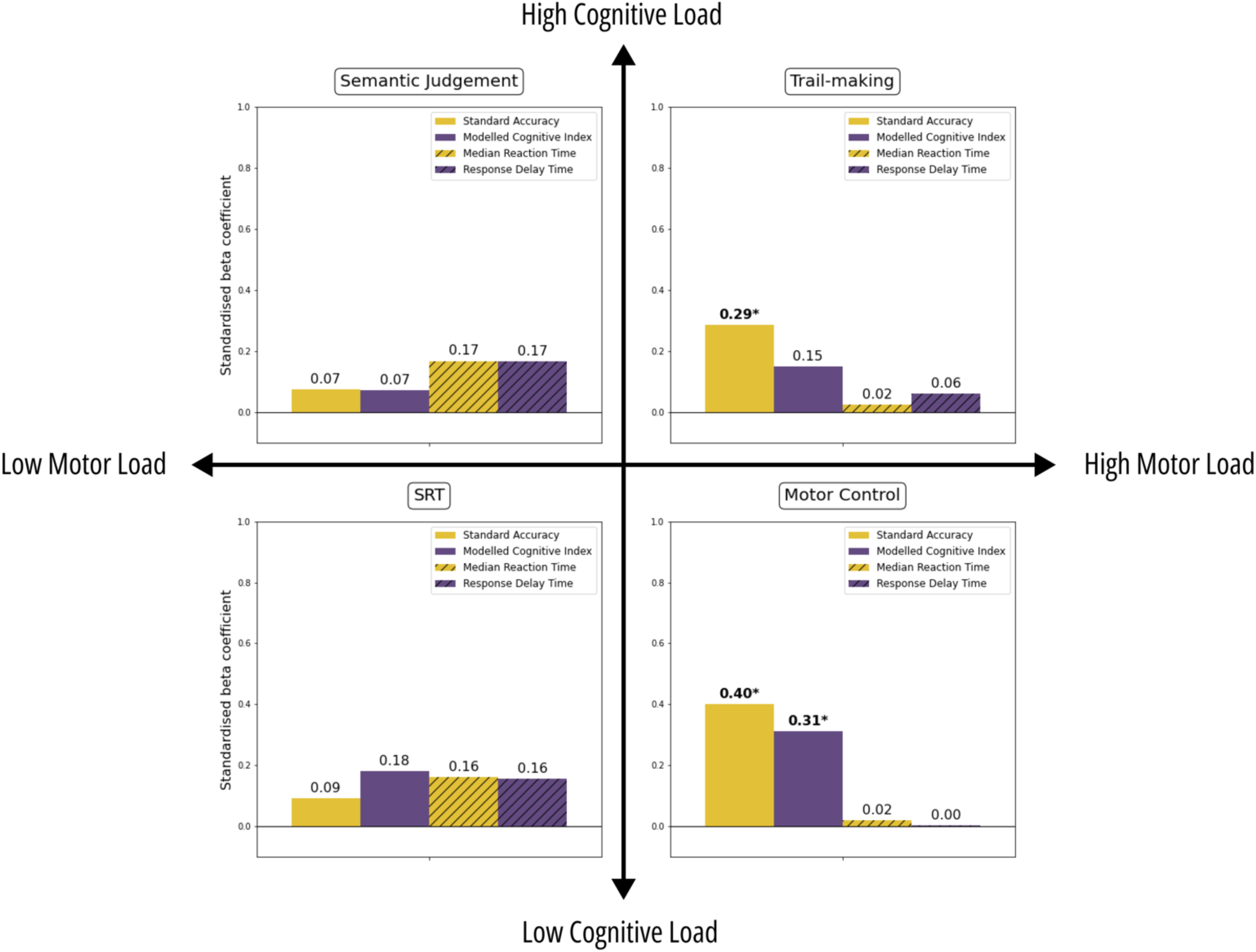
The effect of using an impaired hand when completing the self-administered cognitive tasks in four representative tasks with variable motor and cognitive loadings. The absolute strength of the effect is shown in standardised beta coefficients. Modelled metrics are shown in purple (Cognitive Index= solid purple; Response Delay Time= hatched purple), and standard metrics are shown in amber (Standard accuracy= solid amber; median reaction time in milliseconds= hatched amber). Asterisk in bold denotes significant effect of hand-motor impairment after FDR correction for the four measures within each task. Significant effects indicate worse/slower performance when using an impaired hand.

In contrast, the modelled Cognitive Index, that was expected to account for hand-motor delay, showed no significant effect of using an impaired hand on the cognitive performance for any of the 18 tasks except for the Motor Control task, a task explicitly designed to assess fine hand motor ability (Figure 3). This finding suggests that the modelled Cognitive Index reflects hand impairment solely in tasks specifically designed to measure it and remains unconfounded in tasks targeting particular cognitive processes.

### Modelled Cognitive Index shows a stronger relationship with routinely-used clinical pen-and-paper assessment and functional impairment post-stroke

An overall global factor (G) summarising the performance across all tasks was estimated using principal component analysis (PCA). The principal factor explaining the largest proportion of variance was extracted separately for both the standard accuracy metrics (G_Standard Accuracy_) and the modelled Cognitive Indexes (G_Modelled Cognitive Index_) (see Method). These global cognitive scores were then compared to MoCA scores, a pen-and-paper cognitive screening test, administered by a clinician and therefore expected to be less confounded by hand-motor impairments. The analysis was carried out separately at different phases post-stroke to capture heterogeneity in rates of recovery. Across all stages of stroke recovery, the modelled Cognitive Index exhibited a stronger association with pen-and-paper test scores, explaining, on average, 21% more variance as measured by R-squared (Figure 4).

**Figure 4.**
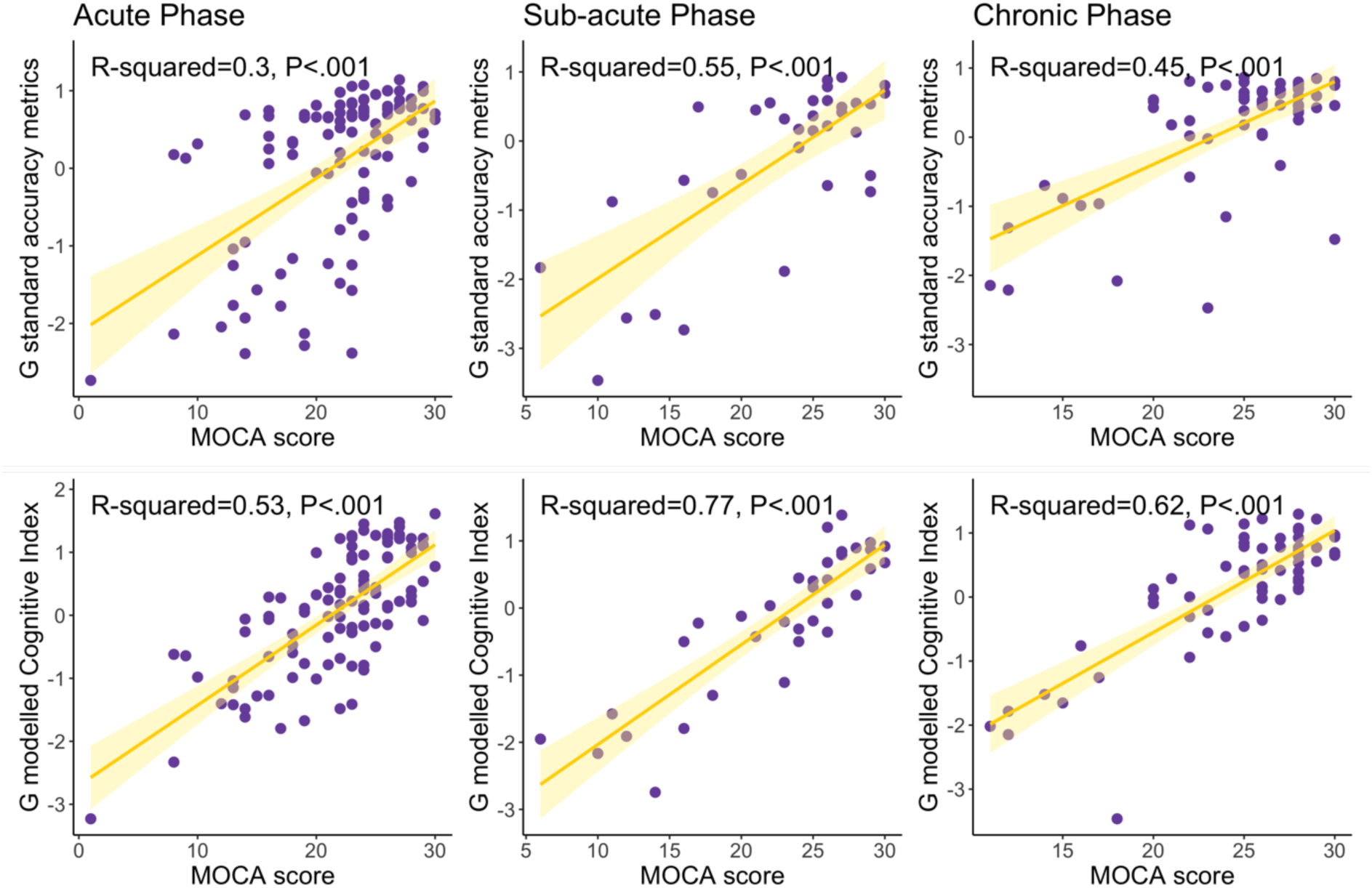
Global cognitive performance derived from the first component of a PCA analysis is related to global performance on a pen-and-paper clinician administered test (MoCA). **Top panel.** Correlation between global standard accuracy metrics and MoCA scores across the acute, sub-acute and chronic phases of the stroke recovery. **Bottom panel.** Correlation between the modelled Cognitive Index, accounting for motoric impairments, across the three phases of stroke reocvery. P-values are uncorrected.

The global measures of cognition were also correlated with functional outcomes post-stroke (IADL scores) related to quality of life (Supplementary Figure 5). These associations were stronger for Cognitive Index during the sub-acute and chronic phases of stroke recovery.

### The modelled Cognitive Index shows stronger relationship with cerebrovascular brain injury load compared with standard scores

The global cognitive performance was related to measures of cerebrovascular brain injury, as determined from stroke lesion and white matter hyperintensity volume, to examine the biological validity of the results. Compared with the standard accuracy metrics, the modelled Cognitive Index showed more significant brain-behaviour associations (p<.05, FDR-correct, Figure 5). These findings indicated that the Cognitive Index is likely to explain more of the variability in cerebrovascular brain injury load, when compared to standard accuracy scores, improving the precision with which cognition can be measured.

**Figure 5.**
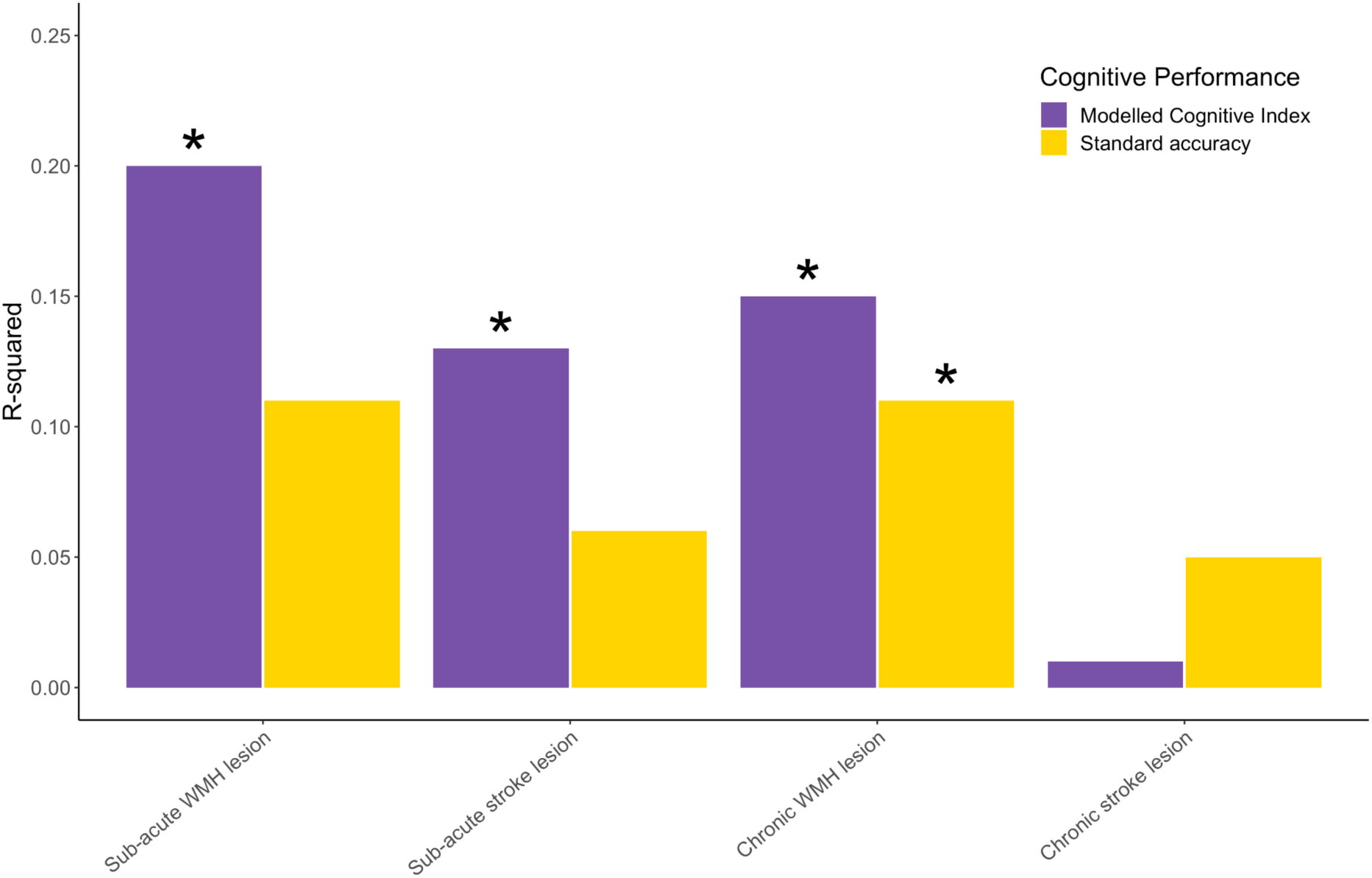
Global cognitive performance estimated via PCA analysis was related to cerebrovascular disease load. Correlations between global cognitive metrics and brain imaging metrics are shown across the sub-acute and chronic phases of stroke where concurrent imaging was available. The Modelled Cognitive Index showed more significant associations with cerebrovascular disease load compared with standard accuracy metrics. Asterisk denotes *P*<.05, FDR-correct.

### Multivariate analyses reveals intuitive relationships between cerebrovascular brain injury load estimates and the modelled cognitive metrics

Canonical Correlation Analyses (CCA) were conducted separately for the sub-acute and chronic stages post-stroke, to examine the many-to-many relationship between the Modelled Cognitive Index derived from all 18 cognitive tasks and measures of cerebrovascular brain injury. Significant multivariate relationships were observed (R^2^=0.49-0.77, P<.001) during both the sub-acute and chronic stages of stroke recovery (see Figure 6), indicating that the Modelled Cognitive metrics and imaging co-vary across all phases of stroke recovery.

**Figure 6.**
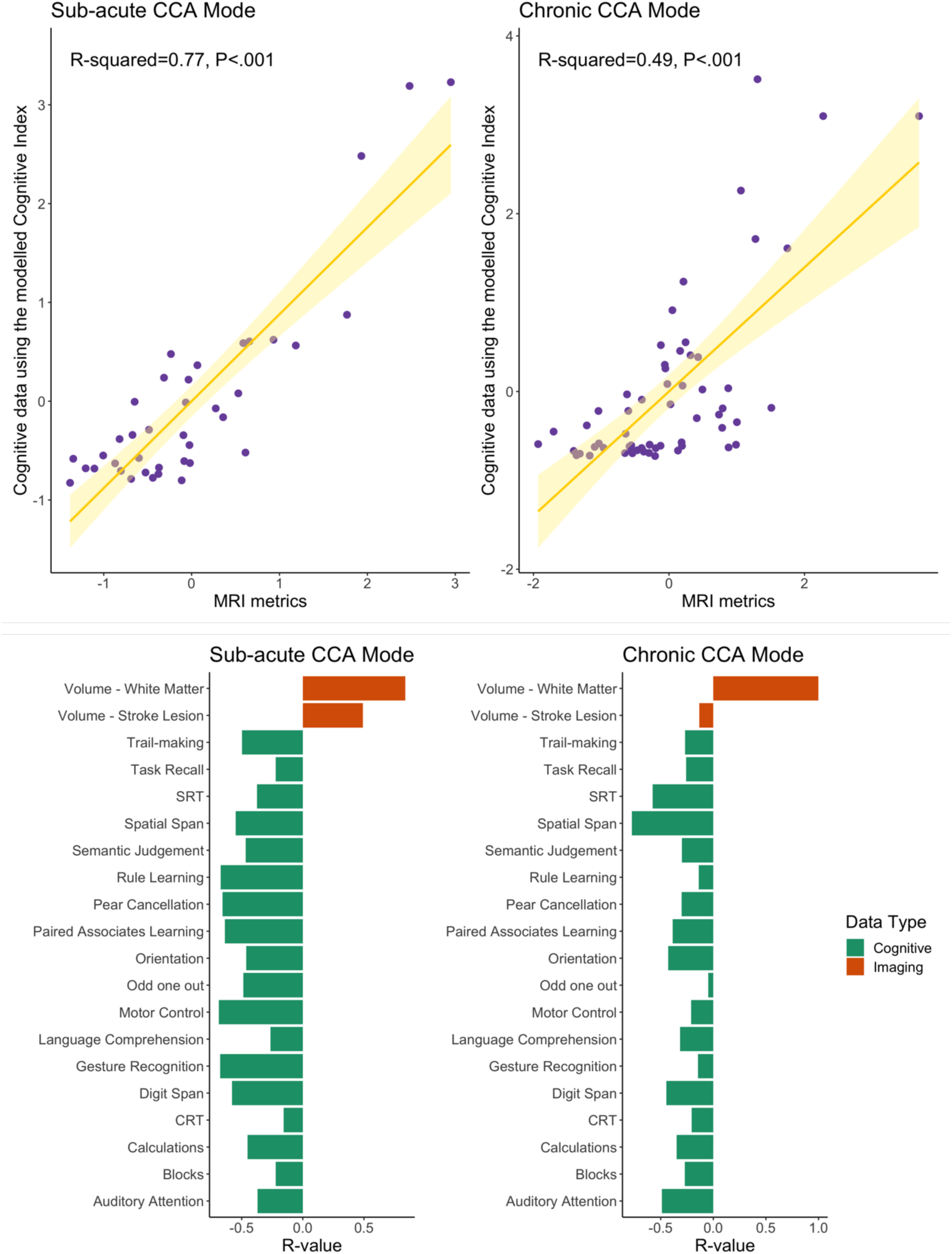
Results of the Canonical Correlation Analysis. **Upper.** The output of the CCA analysis consisted of pairs of canonical variates, which represent individual subject weights that are derived from the cognitive tasks (modelled via the Cognitive Index) with brain imaging measures. The strength of the correlation between the canonical variates indicated whether the mode of population variation is similar to both the pattern of cognition and that of imaging. **Lower**. Back projected r values indicating contribution of the cognitive tasks (green) and brain imaging metric (amber) to the two significant modes derived from the canonical correlation analysis.

More specifically, the CCA revealed that during the sub-acute stage, the combined effect of increased white matter damage and stroke lesion volume was associated with poorer performance across all domains (Figure 6). During the chronic stage, the CCA identified that white matter lesion damage had a stronger loading on tasks that assessed attention, processing speed, and memory (Figure 6). These results are in line with previous literature and provide biological validity for computationally disentangling motor and cognitive contributions to cognitive scores.

## Discussion

The current study evaluated a novel application of a mathematical framework that leverages trial-by-trial variability in self-administered digital tasks to mitigate the confounding effects of motor impairment. The clinical utility and validity of this framework were evaluated in stroke survivors, given their distinctive combination of motor and cognitive deficits, as well as the wide spectrum of cognitive impairments they experience^17^. The model was tested on 18 distinct tasks, which assessed language ability, memory, attention, executive function, reasoning and processing speed, relevant to a broad spectrum of neurological conditions beyond stroke.

The model-derived Cognitive Index provides a more interpretable and intuitive relationship with demographic and clinical factors, compared with standard accuracy metrics. Evidence from a wide range of behavioural domains indicates that cognitive performance, as measured by the modelled Cognitive Index, is not confounded by motor impairments when patients use an impaired hand to select responses. In contrast, cognitive deficits estimated using standard accuracy and/or reaction time metrics tend to be overestimated in the presence of motor impairments, particularly for tasks with high motor demands.

Furthermore, the added clinical utility of the modelled Cognitive Index was demonstrated through its stronger associations with clinical pen-and-paper cognitive assessments (e.g., MoCA) and functional outcomes post-stroke (e.g., IADL), compared to standard accuracy metrics. Finally, the biological validity of the modelled Cognitive Index was established through stronger correlations with neuroimaging-derived metrics of brain injury. Individual tasks also correlated intuitively with specific brain imaging metrics, in line with previous literature^23,24^. Collectively, these findings provide robust evidence for the improved clinical utility and validity of the modelled performance metrics within neurological conditions characterised by co-occurring motor and cognitive deficits.

The afore-mentioned results have broad implications for digital health diagnosis and monitoring that facilitate more efficient and cost-effective means of patient assessments. Whilst an increasing number of remote assessments are being developed and introduced to the market, thus far the confounding effect of motor impairment has been typically dealt with as an exclusion criteria and not directly mitigated^11^. Here, we show that patients with motor impairments need not be excluded from remote clinical testing, by providing a framework that mitigates the effect of those confounding factors. This advancement is particularly beneficial for a wide range of clinical patient populations with co-occurring motor and cognitive deficits, who often experience reduced mobility and independence. Specifically, patients with motor deficits as a result of tremor (e.g., Parkinson’s disease, Lewy body dementia) or paralysis (e.g., traumatic brain injury, cerebral palsy, spinal cord injury, brain tumours, multiple sclerosis, encephalitis, neurodegenerative conditions) would benefit from this inclusive approach to remote assessment. Beyond expanding access to previously underrepresented populations, the framework also enhances the precision and granularity of cognitive assessments, by integrating measures of accuracy with reaction time, and trial difficulty into a unified cognitive outcome which we termed Cognitive Index.

Given the heterogeneity in stroke recovery and its multivariate recovery trajectories across different cognitive domains it was important to test the validity of the model in a wider range of cognitive domains as well as recovery stages^20,25^. We leveraged this unique participant cohort to examine model performance across all stages of stroke recovery (acute, sub-acute, and chronic) and demonstrate that the modelled Cognitive Index produces consistent and intuitive results across the recovery process.

It is important to consider the current findings in the context of certain limitations. While the modelled Cognitive Index effectively mitigates the confounding effects of motor impairment, its capacity to do so is limited to confounds that influence trial-by-trial variability (central to the design of the computational model) rather than a consistent shift in accuracy across all trials. For instance, if a patient were to perform incorrectly on an overwhelming proportion of trials of a given cognitive task purely due to a severe hand impairment, rather than cognitive deficits per se, the model cannot fully eliminate the confounding effects of this fixed motor deficit as there is little variability in the accuracy of the response. Instead, the model is best suited for cases where patients experience mild to moderate hand impairment that affect trial-by-trial performance but do not entirely prevent task completion. Nevertheless, this limitation is not unique to our modelled cognitive metric presented here, but applies to any metric derived from a self-administered task, and explains why in practice such patients are typically excluded from self-administered assessments.

In addition to the modelled Cognitive Index, the IDoCT framework also generates a metric of Response Delay Time. Our results demonstrate that this metric captures variability related to hand-motor impairment —that is, variability no longer reflected in the modelled Cognitive Index. However, the modelled Response Delay Time may also capture other delay sources such as visual or non-specific processes that influence trial-by-trial variability. For example, if a patient experiences higher order visual processing impairment that leads to slower responses across all trials, this delay would be reflected in the Response Delay Time, even in the absence of motor impairment. Thus, it may be challenging to interpret the precise source of an elevated Response Delay Time in patients with co-morbid sensory-motor impairments. Future work will aim to further refine the IDoCT framework to disambiguate these factors, for example by relating quantitative measures of motor and visual impairment to Response Delay Time.

Neurological disorders are the leading cause of disability worldwide.^26^ Here, we provide novel evidence that computational modelling can enhance the precision of unsupervised digital cognitive assessments which will be beneficial in a wide range of neurological disorders^5,27^. By enabling large-scale data collection in a cost-effective manner—while reducing confounds and improving cognitive domain specificity—the IDoCT framework has the potential to reduce diagnostic bottlenecks, improve accessibility, and support early detection and intervention for a broad spectrum of neurological conditions.

## Data Availability

All data produced in the present study are available upon reasonable request to the authors. The Python and R scripts used in the analysis of the paper are publicly available in our github repository.

https://github.com/dragos-gruia/computational_modelling_of_behaviour_in_stroke

## References

1. Abbadessa G, Brigo F, Clerico M, De Mercanti S, Trojsi F, Tedeschi G, et al. Digital therapeutics in neurology. J Neurol. 2022 Mar;269(3):1209–24.

2. Janik MJ, Urbach DV, van Nieuwenhuizen E, Zhao J, Yellin O, Baccara-Dinet MT, et al. Alirocumab treatment and neurocognitive function according to the CANTAB scale in patients at increased cardiovascular risk: A prospective, randomized, placebo-controlled study. Atherosclerosis. 2021 Aug 1;331:20–7.

3. Campos-Magdaleno M, Leiva D, Pereiro AX, Lojo-Seoane C, Mallo SC, Facal D, et al. Changes in visual memory in mild cognitive impairment: a longitudinal study with CANTAB. Psychol Med. 2021 Oct;51(14):2465–75.

4. Toniolo S, Zhao S, Scholcz A, Amein B, Ganse-Dumrath A, Heslegrave AJ, et al. Relationship of plasma biomarkers to digital cognitive tests in Alzheimer’s disease. Alzheimers Dement (Amst). 2024 Apr 14;16(2):e12590.

5. Gupta AS. Digital Phenotyping in Clinical Neurology. Semin Neurol. 2022 Feb;42(01):048–59.

6. The Stroke Association. Priorities in stroke rehabilitation and long-term care. 2021. Available from: https://www.stroke.org.uk/sites/default/files/research/priorities_in_stroke_rehabilitation_and_lon-term_care.pdf

7. Bloem BR, Dorsey ER, Okun MS. The coronavirus disease 2019 crisis as catalyst for telemedicine for chronic neurological disorders. JAMA neurology. 2020;77(8):927–8.

8. Cai Z, Giunchiglia V, Street R, Giovane MD, Lu K, Popham M, et al. Online46: online cognitive assessments in elderly cohorts-the British 1946 birth cohort case study. medRxiv. 2024;2024–09.

9. Lemmens J, Ferdinand S, Vandenbroucke A, Ilsbroukx S, Kos D. Dual-task cost in people with multiple sclerosis: A case–control study. British Journal of Occupational Therapy. 2018 Jul 1;81(7):384–92.

10. Low E, Crewther SG, Ong B, Perre D, Wijeratne T. Compromised Motor Dexterity Confounds Processing Speed Task Outcomes in Stroke Patients. Front Neurol. 2017 Sep 21. Available from: https://www.frontiersin.org/journals/neurology/articles/10.3389/fneur.2017.004 84/full

11. Chen C, Leys D, Esquenazi A. The interaction between neuropsychological and motor deficits in patients after stroke. Neurology. 2013 Jan 15;80(3_supplement_2):S27–34.

12. Demeyere N, Haupt M, Webb SS, Strobel L, Milosevich ET, Moore MJ, et al. Introducing the tablet-based Oxford Cognitive Screen-Plus (OCS-Plus) as an assessment tool for subtle cognitive impairments. Sci Rep. 2021 Apr 12;11(1):8000.

13. Giunchiglia V, Gruia DC, Lerede A, Trender W, Hellyer P, Hampshire A. An iterative approach for estimating domain-specific cognitive abilities from large scale online cognitive data. npj Digital Medicine. 2024;7(1):328.

14. Giunchiglia V, Curtis S, Smith S, Allen N, Hampshire A. Neural correlates of cognitive ability and visuo-motor speed: validation of IDoCT on UK Biobank Data. Imaging Neuroscience. 2024;2:1–25.

15. Gruia DC, Giunchiglia V, Coghlan A, Brook S, Banerjee S, Kwan J, et al. Online monitoring technology for deep phenotyping of cognitive impairment after stroke. medRxiv. 2024;2024–09.

16. Gruia DC, Trender W, Hellyer P, Banerjee S, Kwan J, Zetterberg H, et al. IC3 protocol: a longitudinal observational study of cognition after stroke using novel digital health technology. BMJ Open. 2023 Nov 1;13(11):e076653.

17. Demeyere N, Riddoch MJ, Slavkova ED, Jones K, Reckless I, Mathieson P, et al. Domain-specific versus generalized cognitive screening in acute stroke. J Neurol. 2016 Feb 1;263(2):306–15.

18. Ramsey L, Siegel J, Lang C, Strube M, Shulman G, Corbetta M. Behavioural clusters and predictors of performance during recovery from stroke. Nat Hum Behav. 2017;1:0038.

19. Oba S, Sato M aki, Takemasa I, Monden M, Matsubara K ichi, Ishii S. A Bayesian missing value estimation method for gene expression profile data. Bioinformatics. 2003;19(16):2088–96.

20. Stefaniak JD, Geranmayeh F, Lambon Ralph MA. The multidimensional nature of aphasia recovery post-stroke. Brain. 2022;145(4):1354–67.

21. Hope TMH, Friston K, Price CJ, Leff AP, Rotshtein P, Bowman H. Recovery after stroke: not so proportional after all? Brain. 2019 Jan 1;142(1):15–22.

22. Wang HT, Smallwood J, Mourao-Miranda J, Xia CH, Satterthwaite TD, Bassett DS, et al. Finding the needle in a high-dimensional haystack: Canonical correlation analysis for neuroscientists. NeuroImage. 2020 Aug 1;216:116745.

23. Kinnunen KM, Greenwood R, Powell JH, Leech R, Hawkins PC, Bonnelle V, et al. White matter damage and cognitive impairment after traumatic brain injury. Brain. 2011;134(2):449–63.

24. Sperber C, Gallucci L, Mirman D, Arnold M, Umarova RM. Stroke lesion size – Still a useful biomarker for stroke severity and outcome in times of high-dimensional models. NeuroImage: Clinical. 2023 Jan 1;40:103511.

25. Saa JP, Tse T, Baum C, Cumming T, Josman N, Rose M, et al. Longitudinal evaluation of cognition after stroke–A systematic scoping review. PloS one. 2019;14(8):e0221735.

26. Deuschl G, Beghi E, Fazekas F, Varga T, Christoforidi KA, Sipido E, et al. The burden of neurological diseases in Europe: an analysis for the Global Burden of Disease Study 2017. The Lancet Public Health. 2020;5(10):e551–67.

27. Germine L, Reinecke K, Chaytor NS. Digital neuropsychology: Challenges and opportunities at the intersection of science and software. The Clinical Neuropsychologist. 2019 Feb 17;33(2):271–86.

