## Supplementary Material for "Mitigating the impact of motor impairment on self-administered digital tests in patients with neurological disorders"

### Supplementary Methods

#### MRI data acquisition and pre-processing

Structural MRI data, including high-resolution $1mm^3$ isotropic T1-weighted and T2-FLAIR (Fluid Attenuated Inversion Recovery) sequences were acquired using a Siemens 3T scanner at three and twelve months after stroke. T1-weighted sequences were obtained using the following parameters: repetition time (TR)=2300ms, echo time (TE)=2.98ms, flip angle (FA)=9°, field of view (FOV)=265x265mm, 160 slices. T2-FLAIR acquisition parameters were: TR=5000ms, TE=395ms, FA=120°, FOV=250x250mm, 160 slices.

Lesion masks and white matter hyperintensities (WMH) masks were manually delineated on the T2-FLAIR images guided by the clinical diffusion weighted imaging (DWI) obtained acutely after the stroke. All masks were created by trained research teams and checked by experienced neurologist (FG). Non-brain voxels were removed from T1 and T2-FLAIR images using the Brain Extraction Tool (BET) from FMRIB's Software Library (FSL), using robust brain centre estimation and a fractional intensity threshold of 0.5. T2-FLAIR brain images, along with lesion and WMH masks, were registered to T1 space with 6 degrees of freedom (DOF), using FMRIB's Linear Image Registration Tool (FLIRT). Subsequently, T1 brain images and masks were registered to the standard Montreal Neurological Institute (MNI) 1mm brain template using 12 DOF and trilinear interpolation. An inverted lesion mask was included in the registration to down-weight the influence of the lesion in the registration and thus minimize distortions associated with the registration of lesioned tissue. Volumetric measures of the standard-space lesion and WMH masks were calculated for each subject using FSL. The lesion overlap maps are presented in Supplementary Figure 9, separately for stroke lesion and white matter hyperintensities.

### Supplementary Results

**Supplementary Figure 1**: **Difficulty scale assigned to each of the 18 task.** Y axis shows scaled difficulty of each trial type in arbitrary units derived from the computational model. X axis shows the trial types per task. **Trail-Making Task.** The task includes 14 “Circles” and “Stars” trials that do not require switching, and 14 “Switching” trials that require participants to alternate between categories. It is observed that, generally, the “Switching” trials are more challenging than the “Circles-only” and “Stars-only” trials. The first trial is typically the most difficult, regardless of whether switching is required. **SRT (Simple Reaction Time) and Auditory Attention Tasks.** Those tasks are modelled using the inter-stimulus interval (ISI) between the current and previous trial. Trials with consistent ISI are generally easier with faster and more accurate responses. In contrast, variable ISI leads to slower responses and increased errors. **Rule Learning Task.** The task consists of eight sequential “rule changes” (labelled 1 to 8) that participants must figure out. The rules become progressively more challenging, with the hardest rules presented towards the end of the task. **Motor Control Task.** Difficulty is modelled based on the distance (in pixels) between the ‘current’ target and the previously seen target. Larger distances require more complex hand movements, leading to slower response times

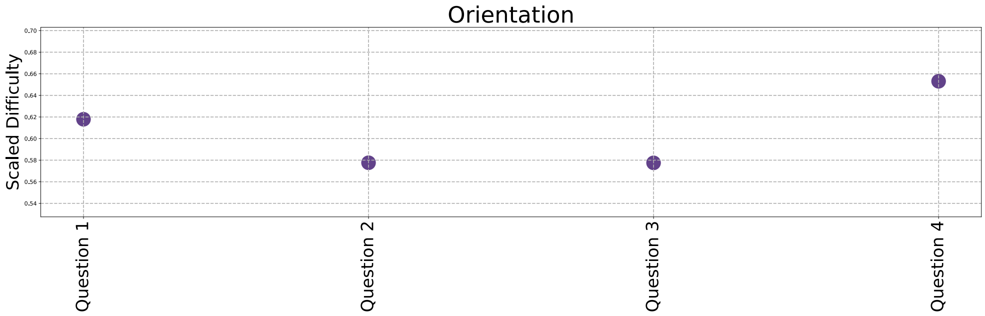

Orientation
Scaled Difficulty
Question 1
Question 2
Question 3
Question 4


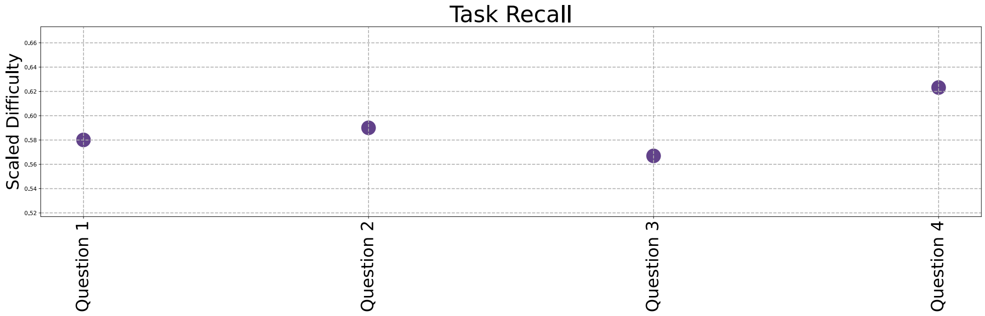

Task Recall
Scaled Difficulty
Question 1
Question 2
Question 3
Question 4


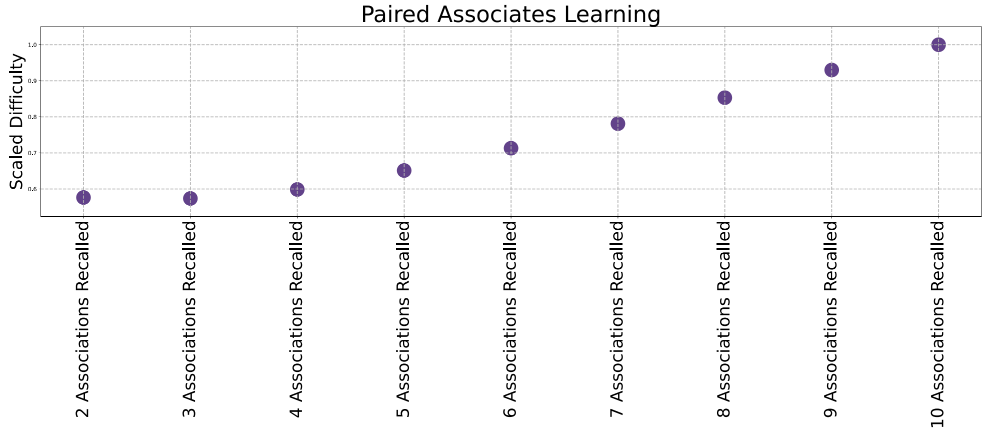

Paired Associates Learning
Scaled Difficulty
2 Associations Recalled
3 Associations Recalled
4 Associations Recalled
5 Associations Recalled
6 Associations Recalled
7 Associations Recalled
8 Associations Recalled
9 Associations Recalled
10 Associations Recalled


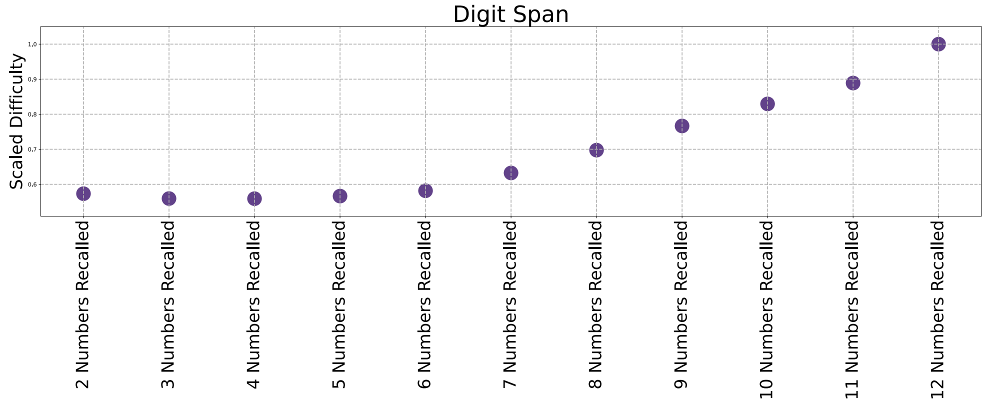

Digit Span
Scaled Difficulty
2 Numbers Recalled
3 Numbers Recalled
4 Numbers Recalled
5 Numbers Recalled
6 Numbers Recalled
7 Numbers Recalled
8 Numbers Recalled
9 Numbers Recalled
10 Numbers Recalled
11 Numbers Recalled
12 Numbers Recalled

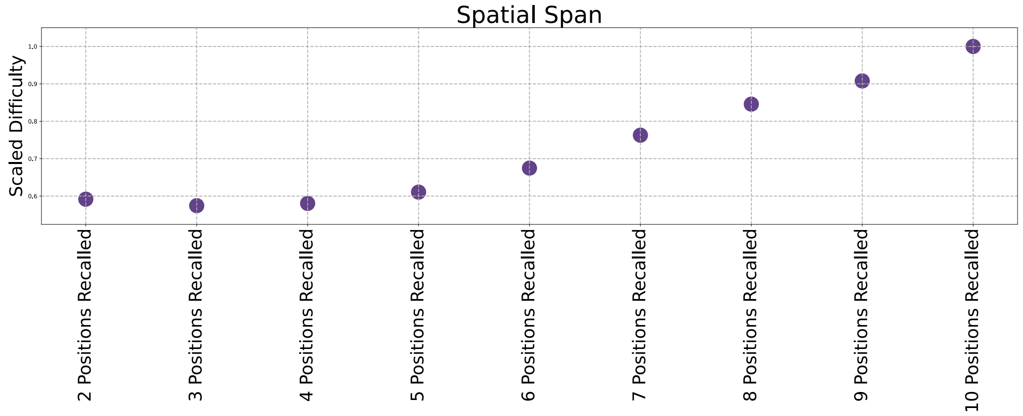
Spatial Span
Scaled Difficulty
2 Positions Recalled
3 Positions Recalled
4 Positions Recalled
5 Positions Recalled
6 Positions Recalled
7 Positions Recalled
8 Positions Recalled
9 Positions Recalled
10 Positions Recalled

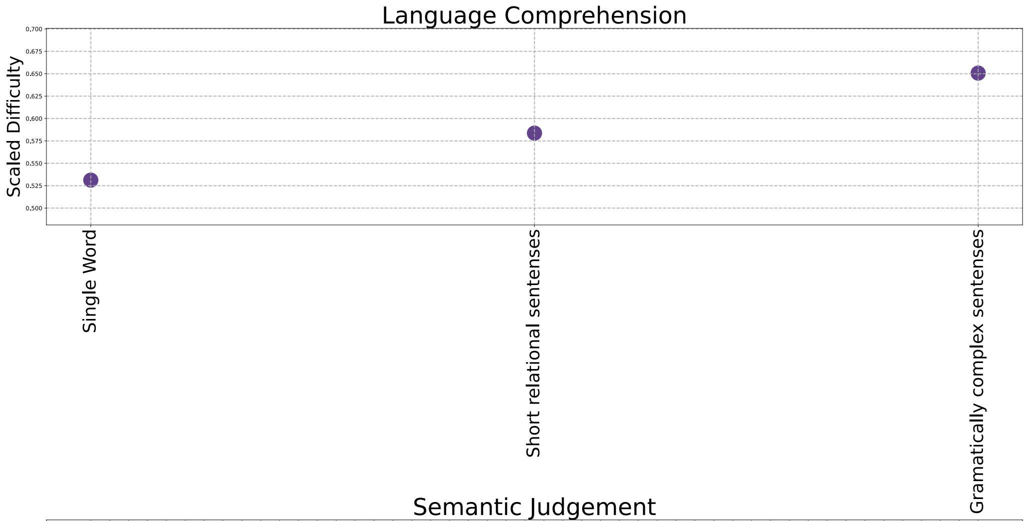
Language Comprehension
Scaled Difficulty
Single Word
Short relational sentenses
Gramatically complex sentenses

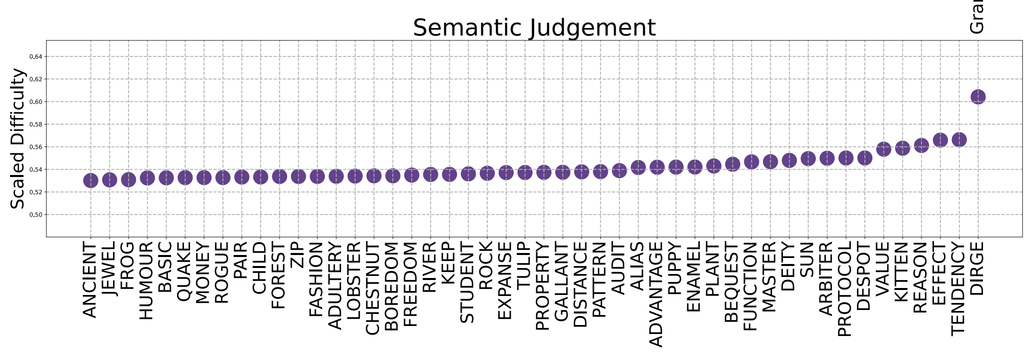
Semantic Judgement
Scaled Difficulty
ANCIENT
JEWEL
FROG
HUMOUR
BASIC
QUAKE
MONEY
ROGUE
PAIR
CHILD
FOREST
ZIP
FASHION
ADULTERY
LOBSTER
CHESTNUT
BOREDOM
FREEDOM
RIVER
KEEP
STUDENT
ROCK
EXPANSE
TULIP
PROPERTY
GALLANT
DISTANCE
PATTERN
AUDIT
ALIAS
ADVANTAGE
PUPPY
ENAMEL
PLANT
BEQUEST
FUNCTION
MASTER
DEITY
SUN
ARBITER
PROTOCOL
DESPOT
VALUE
KITTEN
REASON
EFFECT
TENDENCY
DIRGE

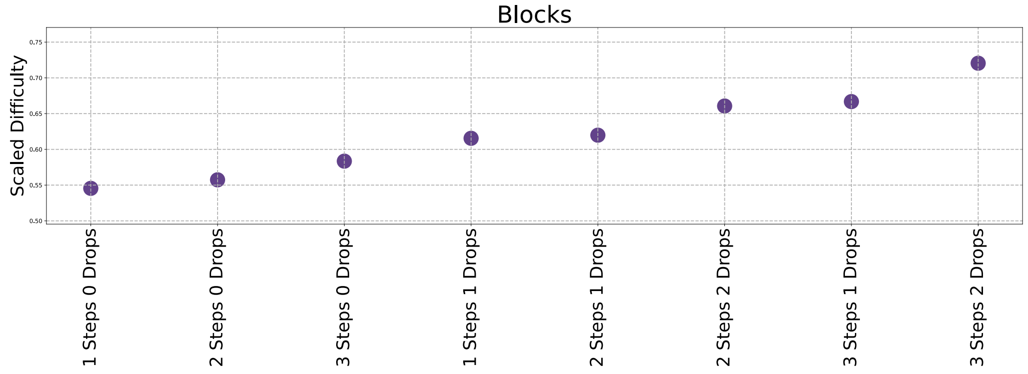
Blocks
Scaled Difficulty
1 Steps 0 Drops
2 Steps 0 Drops
3 Steps 0 Drops
1 Steps 1 Drops
2 Steps 1 Drops
2 Steps 2 Drops
3 Steps 1 Drops
3 Steps 2 Drops

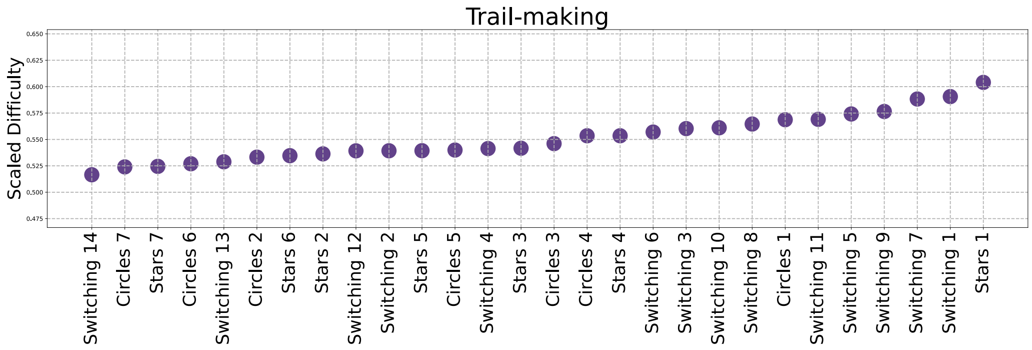
Trail-making
Scaled Difficulty
Switching 14
Circles 7
Stars 7
Circles 6
Switching 13
Circles 2
Stars 6
Stars 2
Switching 12
Switching 2
Stars 5
Circles 5
Switching 4
Stars 3
Circles 3
Circles 4
Stars 4
Switching 6
Switching 3
Switching 10
Switching 8
Circles 1
Switching 11
Switching 5
Switching 9
Switching 7
Switching 1
Stars 1

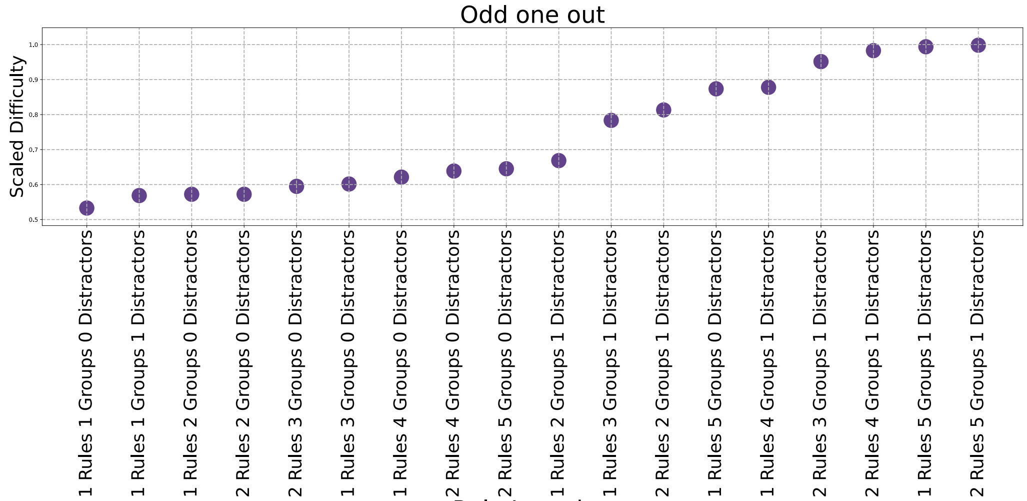
Odd one out
Scaled Difficulty
1 Rules 1 Groups 0 Distractors
1 Rules 1 Groups 1 Distractors
1 Rules 2 Groups 0 Distractors
2 Rules 2 Groups 0 Distractors
2 Rules 3 Groups 0 Distractors
1 Rules 3 Groups 0 Distractors
1 Rules 4 Groups 0 Distractors
2 Rules 4 Groups 0 Distractors
2 Rules 5 Groups 0 Distractors
1 Rules 2 Groups 1 Distractors
1 Rules 3 Groups 1 Distractors
2 Rules 2 Groups 1 Distractors
1 Rules 5 Groups 0 Distractors
1 Rules 4 Groups 1 Distractors
2 Rules 3 Groups 1 Distractors
2 Rules 4 Groups 1 Distractors
1 Rules 5 Groups 1 Distractors
2 Rules 5 Groups 1 Distractors

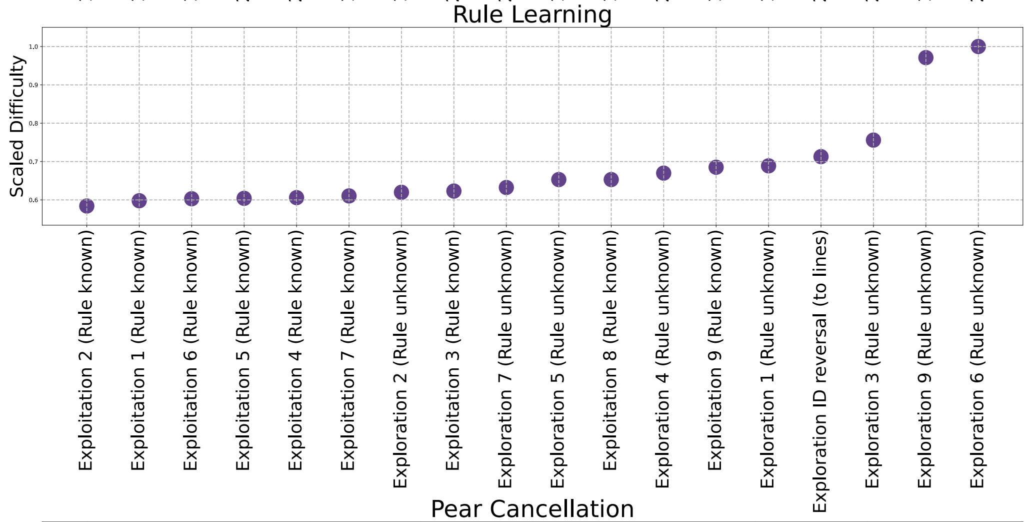
Rule Learning
Scaled Difficulty
Exploitation 2 (Rule known)
Exploitation 1 (Rule known)
Exploitation 6 (Rule known)
Exploitation 5 (Rule known)
Exploitation 4 (Rule known)
Exploitation 7 (Rule known)
Exploration 2 (Rule unknown)
Exploitation 3 (Rule known)
Exploration 7 (Rule unknown)
Exploration 5 (Rule unknown)
Exploitation 8 (Rule known)
Exploration 4 (Rule unknown)
Exploitation 9 (Rule known)
Exploration 1 (Rule unknown)
Exploration ID reversal (to lines)
Exploration 3 (Rule unknown)
Exploration 9 (Rule unknown)
Exploration 6 (Rule unknown)

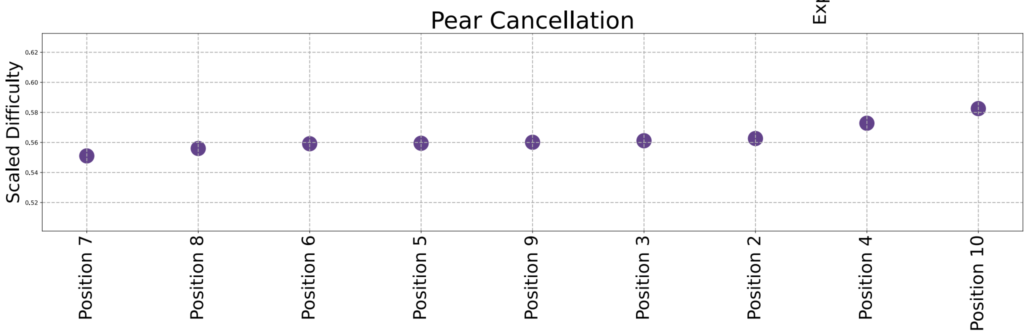
Pear Cancellation
Scaled Difficulty
Position 7
Position 8
Position 6
Position 5
Position 9
Position 3
Position 2
Position 4
Position 10

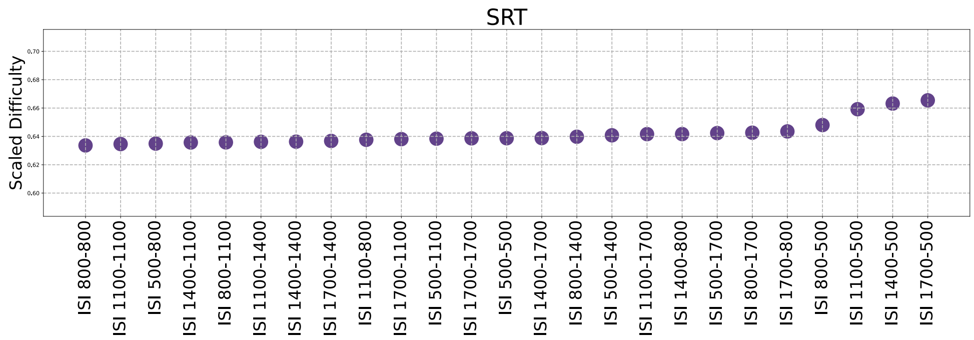
SRT
Scaled Difficulty
ISI 800-800
ISI 1100-1100
ISI 500-800
ISI 1400-1100
ISI 800-1100
ISI 1100-1400
ISI 1400-1400
ISI 1700-1400
ISI 1100-800
ISI 1700-1100
ISI 500-1100
ISI 1700-1700
ISI 500-500
ISI 1400-1700
ISI 800-1400
ISI 500-1400
ISI 1100-1700
ISI 1400-800
ISI 500-1700
ISI 800-1700
ISI 1700-800
ISI 800-500
ISI 1100-500
ISI 1400-500
ISI 1700-500

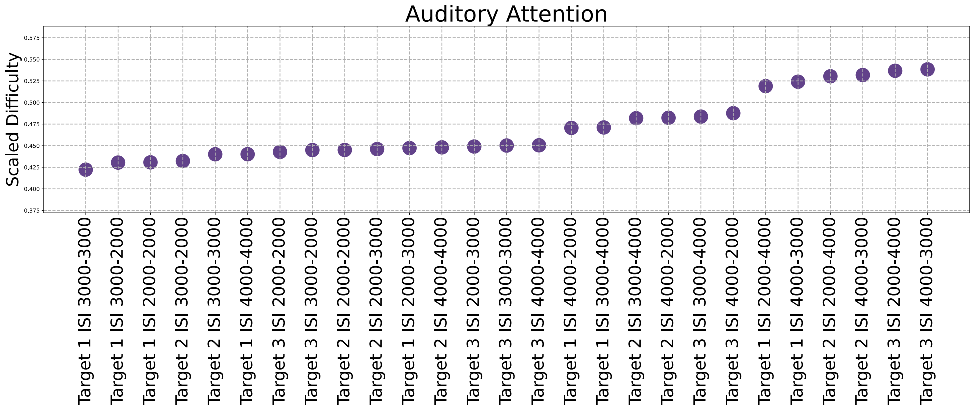
Auditory Attention
Scaled Difficulty
Target 1 ISI 3000-3000
Target 1 ISI 3000-2000
Target 1 ISI 2000-2000
Target 2 ISI 3000-2000
Target 2 ISI 3000-3000
Target 1 ISI 4000-4000
Target 3 ISI 2000-2000
Target 3 ISI 3000-2000
Target 2 ISI 2000-2000
Target 2 ISI 2000-3000
Target 1 ISI 2000-3000
Target 2 ISI 4000-4000
Target 3 ISI 2000-3000
Target 3 ISI 3000-3000
Target 3 ISI 4000-4000
Target 1 ISI 4000-2000
Target 1 ISI 3000-4000
Target 2 ISI 3000-4000
Target 2 ISI 4000-2000
Target 3 ISI 3000-4000
Target 3 ISI 4000-2000
Target 1 ISI 2000-4000
Target 1 ISI 4000-3000
Target 2 ISI 2000-4000
Target 2 ISI 4000-3000
Target 3 ISI 2000-4000
Target 3 ISI 4000-3000

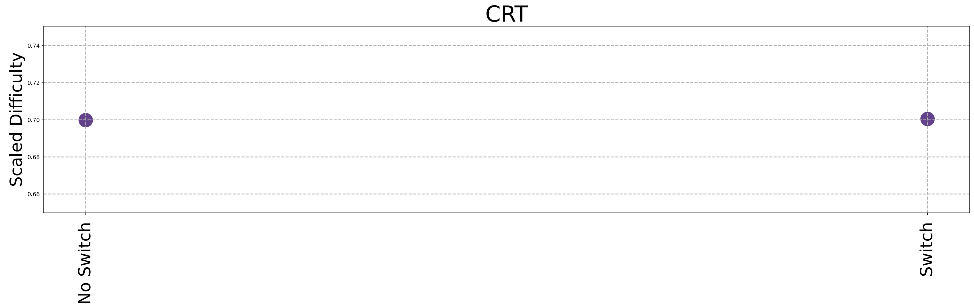
CRT
Scaled Difficulty
No Switch
Switch

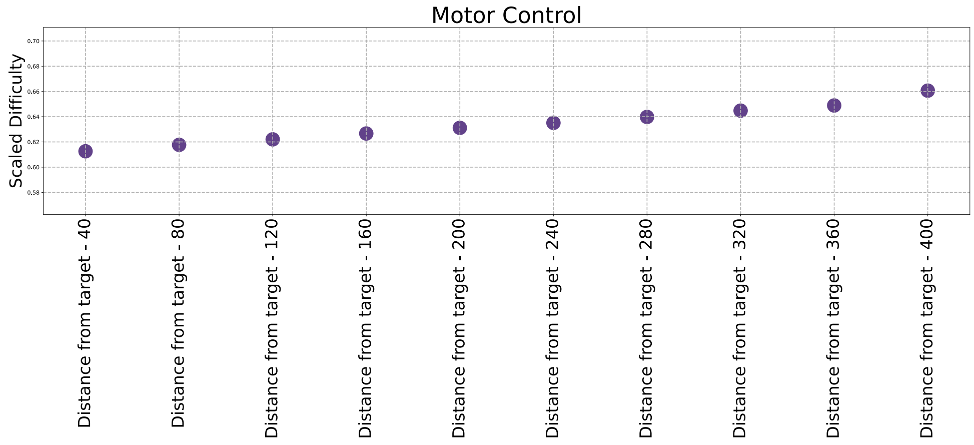
Motor Control
Scaled Difficulty
Distance from target - 40
Distance from target - 80
Distance from target - 120
Distance from target - 160
Distance from target - 200
Distance from target - 240
Distance from target - 280
Distance from target - 320
Distance from target - 360
Distance from target - 400

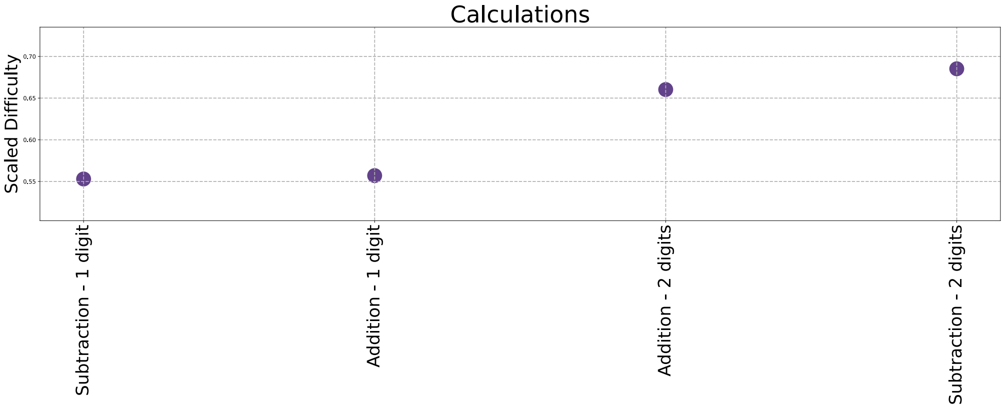
Calculations
Scaled Difficulty
0.70
0.65
0.60
0.55
Subtraction - 1 digit
Addition - 1 digit
Addition - 2 digits
Subtraction - 2 digits

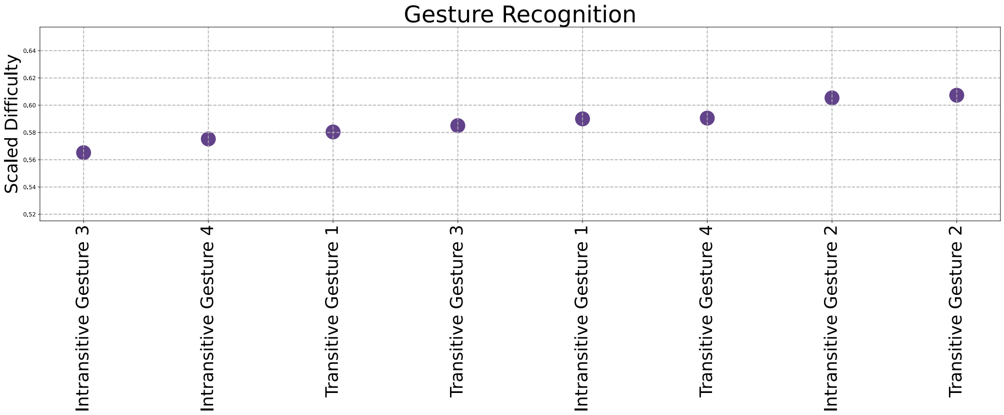
Gesture Recognition
Scaled Difficulty
0.64
0.62
0.60
0.58
0.56
0.54
0.52
Intransitive Gesture 3
Intransitive Gesture 4
Transitive Gesture 1
Transitive Gesture 3
Intransitive Gesture 1
Transitive Gesture 4
Intransitive Gesture 2
Transitive Gesture 2

**Supplementary Figure 2** - Outcome measure distributions for each task. Yellow bars refer to standard raw metrics derived for each task (Standard Accuracy and Median Reaction Time in milliseconds). Purple refers to modelled performance measures (Cognitive Index and Response Delay Time in standard deviation units).

Across all 18 tasks, raw accuracy metrics were predominantly observed to follow a negatively skewed gamma distribution, expected for such clinical research cohort where the patients tend to suffer from mild-moderate impairment levels. In contrast, the modelled Cognitive Index more closely approximated to a Gaussian distribution, thereby mitigating ceiling effects, and showing its ability to capture more variability in cognition. Speed-related measures (e.g., median reaction time and Response Delay Time) typically followed a gamma distribution with a slight positive skew. This is in keeping with speed-based measures capturing a wider dynamic range.

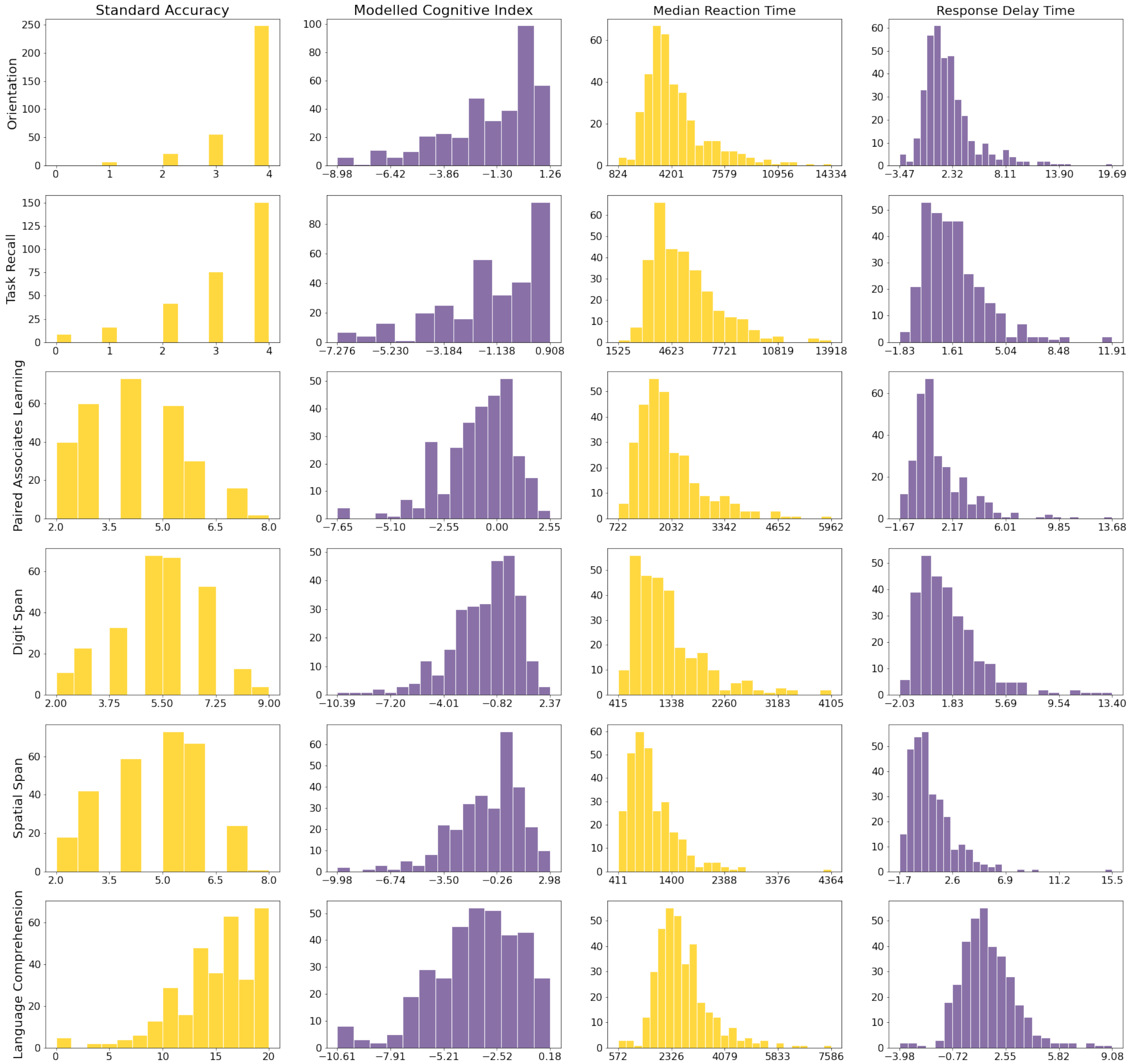

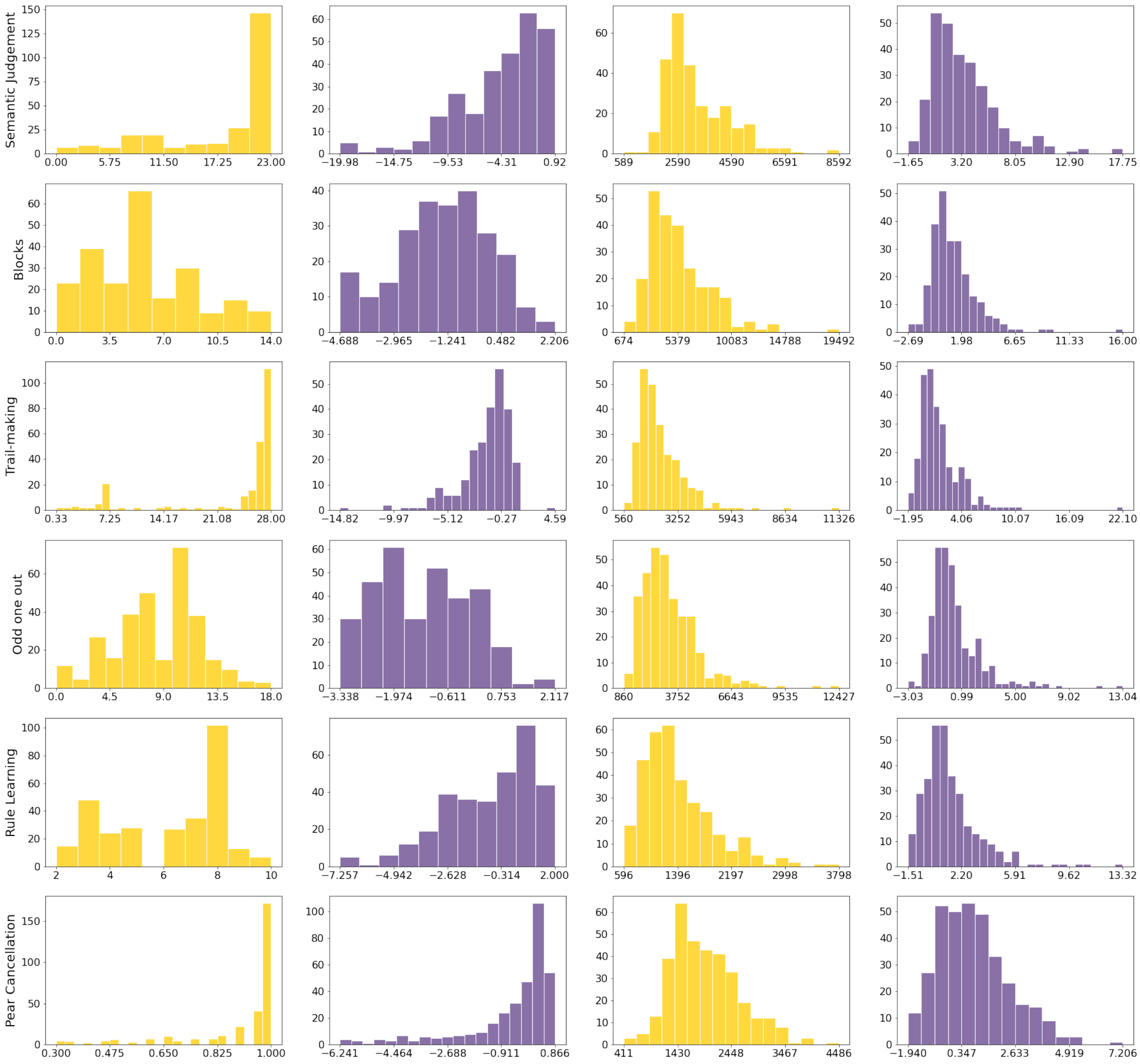
Semantic Judgement
0.00 5.75 11.50 17.25 23.00
−19.98 −14.75 −9.53 −4.31 0.92
589 2590 4590 6591 8592
−1.65 3.20 8.05 12.90 17.75
Blocks
0.0 3.5 7.0 10.5 14.0
−4.688 −2.965 −1.241 0.482 2.206
674 5379 10083 14788 19492
−2.69 1.98 6.65 11.33 16.00
Trail-making
0.33 7.25 14.17 21.08 28.00
−14.82 −9.97 −5.12 −0.27 4.59
560 3252 5943 8634 11326
−1.95 4.06 10.07 16.09 22.10
Odd one out
0.0 4.5 9.0 13.5 18.0
−3.338 −1.974 −0.611 0.753 2.117
860 3752 6643 9535 12427
−3.03 0.99 5.00 9.02 13.04
Rule Learning
2 4 6 8 10
−7.257 −4.942 −2.628 −0.314 2.000
596 1396 2197 2998 3798
−1.51 2.20 5.91 9.62 13.32
Pear Cancellation
0.300 0.475 0.650 0.825 1.000
−6.241 −4.464 −2.688 −0.911 0.866
411 1430 2448 3467 4486
−1.940 0.347 2.633 4.919 7.206

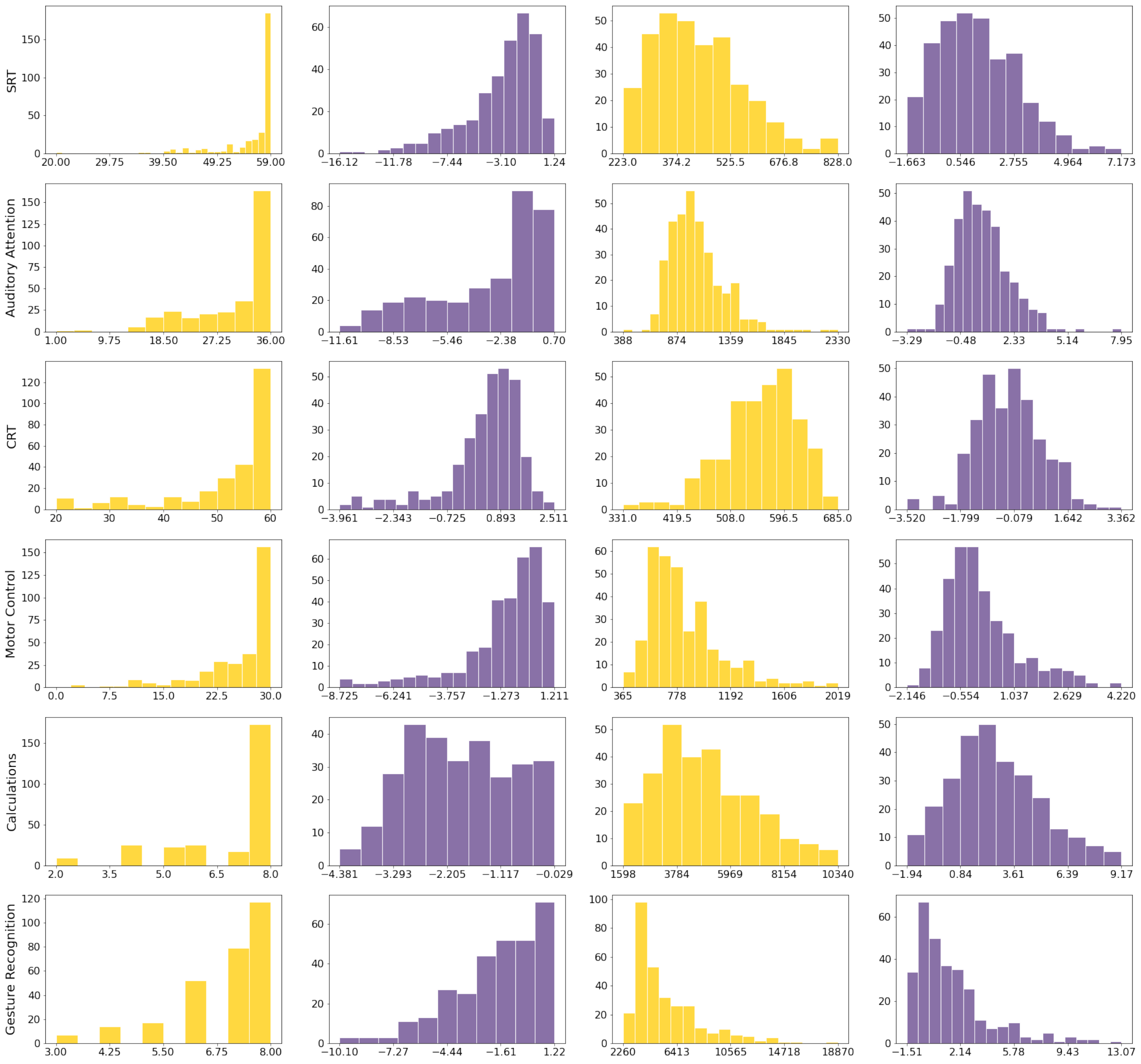

SRT
Auditory Attention
CRT
Motor Control
Calculations
Gesture Recognition

**Supplementary Figure 3 –** Relationship between cognitive scores and demographic and clinical variables. The modelled Cognitive Index is displayed in purple. Standard accuracy scores are shown in yellow. The strength of the association is shown in standard deviation units, along with 95% confidence intervals. The reference value for education refers to patients that were at most educated to Secondary school (≤16 years old, GCSE level).

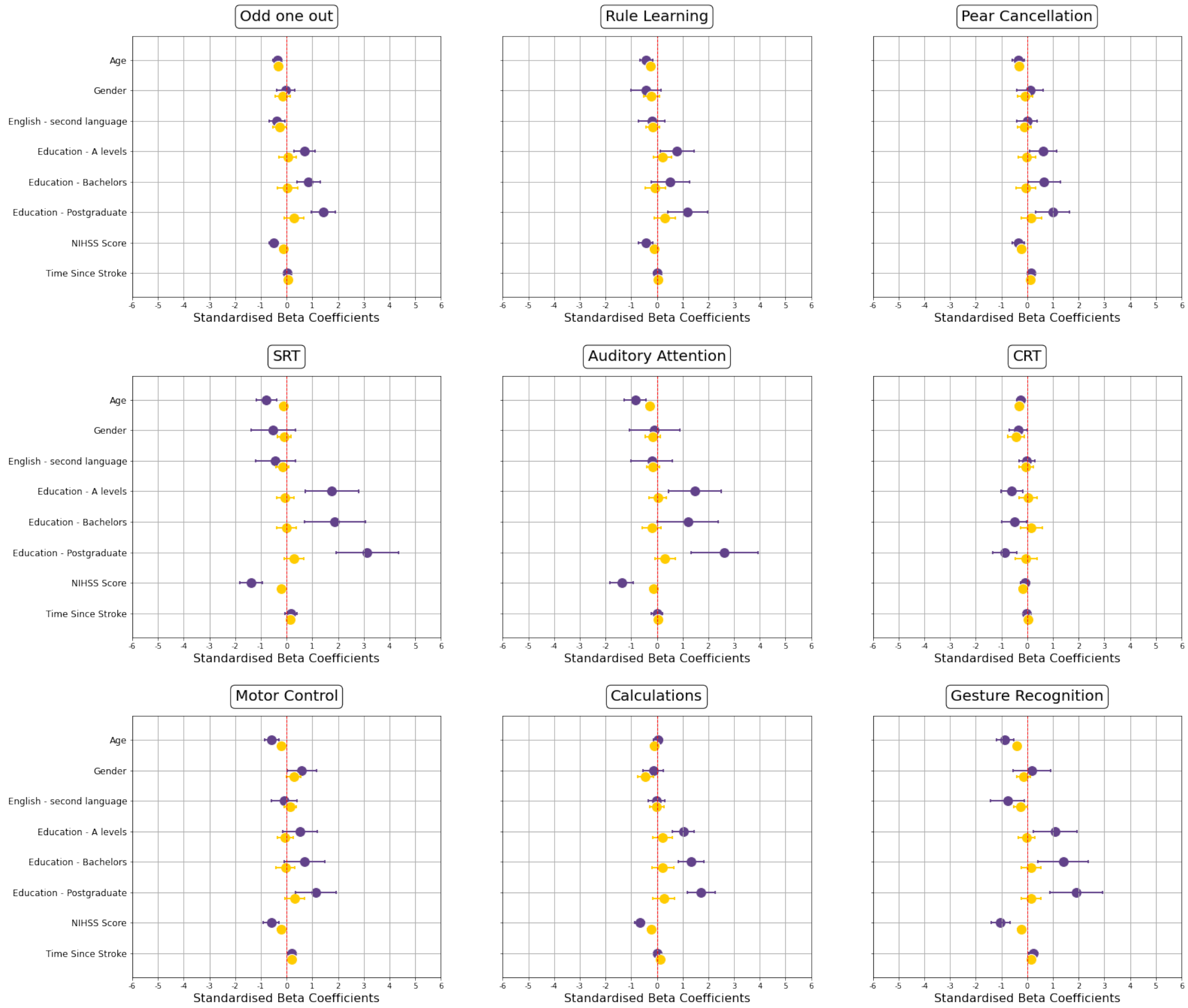

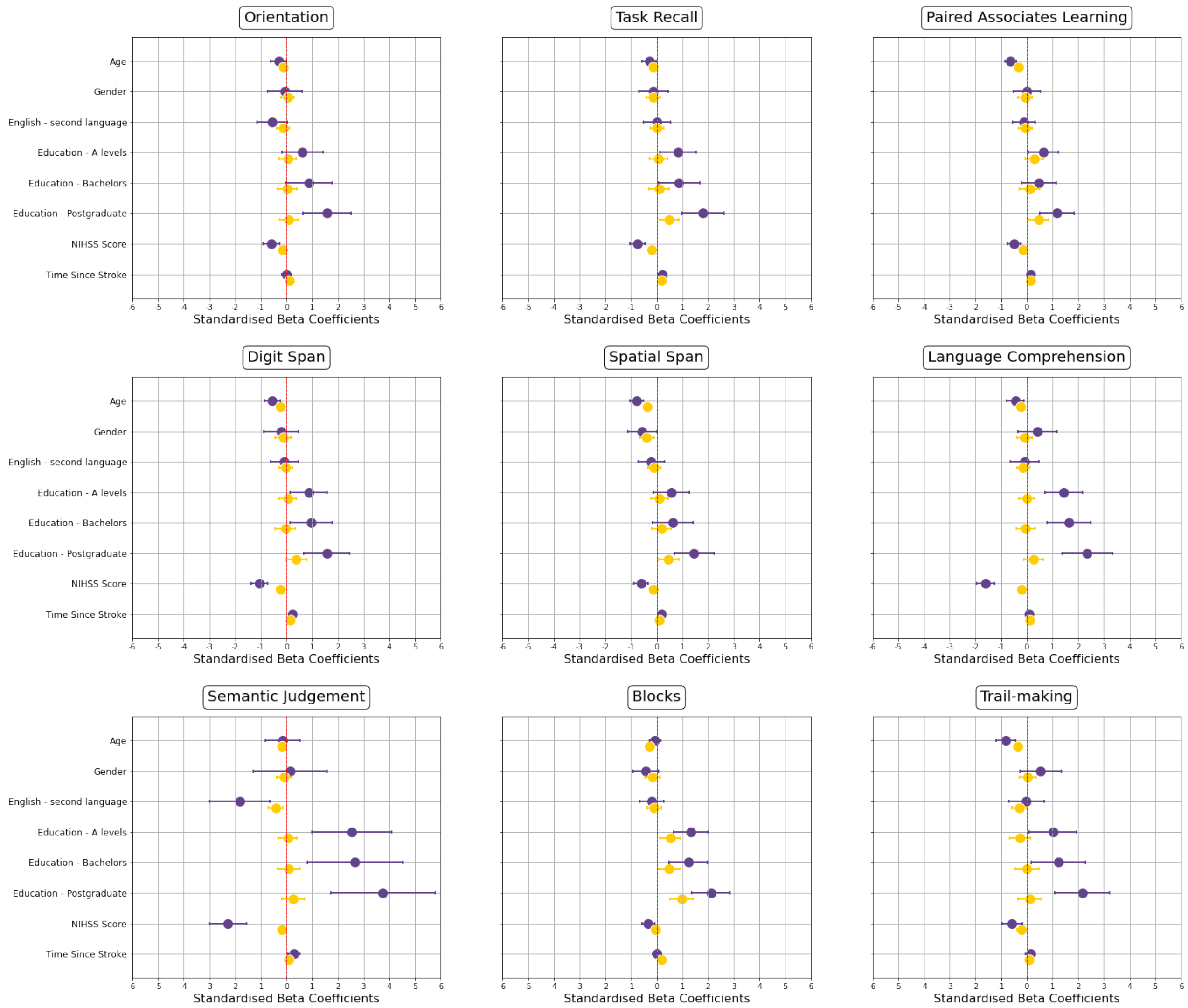

Orientation
Task Recall
Paired Associates Learning
Digit Span
Spatial Span
Language Comprehension
Semantic Judgement
Blocks
Trail-making
Age
Gender
English - second language
Education - A levels
Education - Bachelors
Education - Postgraduate
NIHSS Score
Time Since Stroke
Standardised Beta Coefficients

**Supplementary Figure 4 –** The effect of using an impaired hand when completing the self-administered cognitive tasks. The absolute strength of the effect is shown in standardised beta coefficients. Modelled metrics are shown in purple (Cognitive Index= solid purple; Response Delay Time= hatched purple), and standard metrics are shown in amber (Standard accuracy= solid amber; median reaction time in milliseconds (hatched amber). Asterisk denotes significant effect of hand-motor impairment after FDR correction for the four performance metrics within each task. Significant effects indicate worse/slower performance when using an impaired hand.

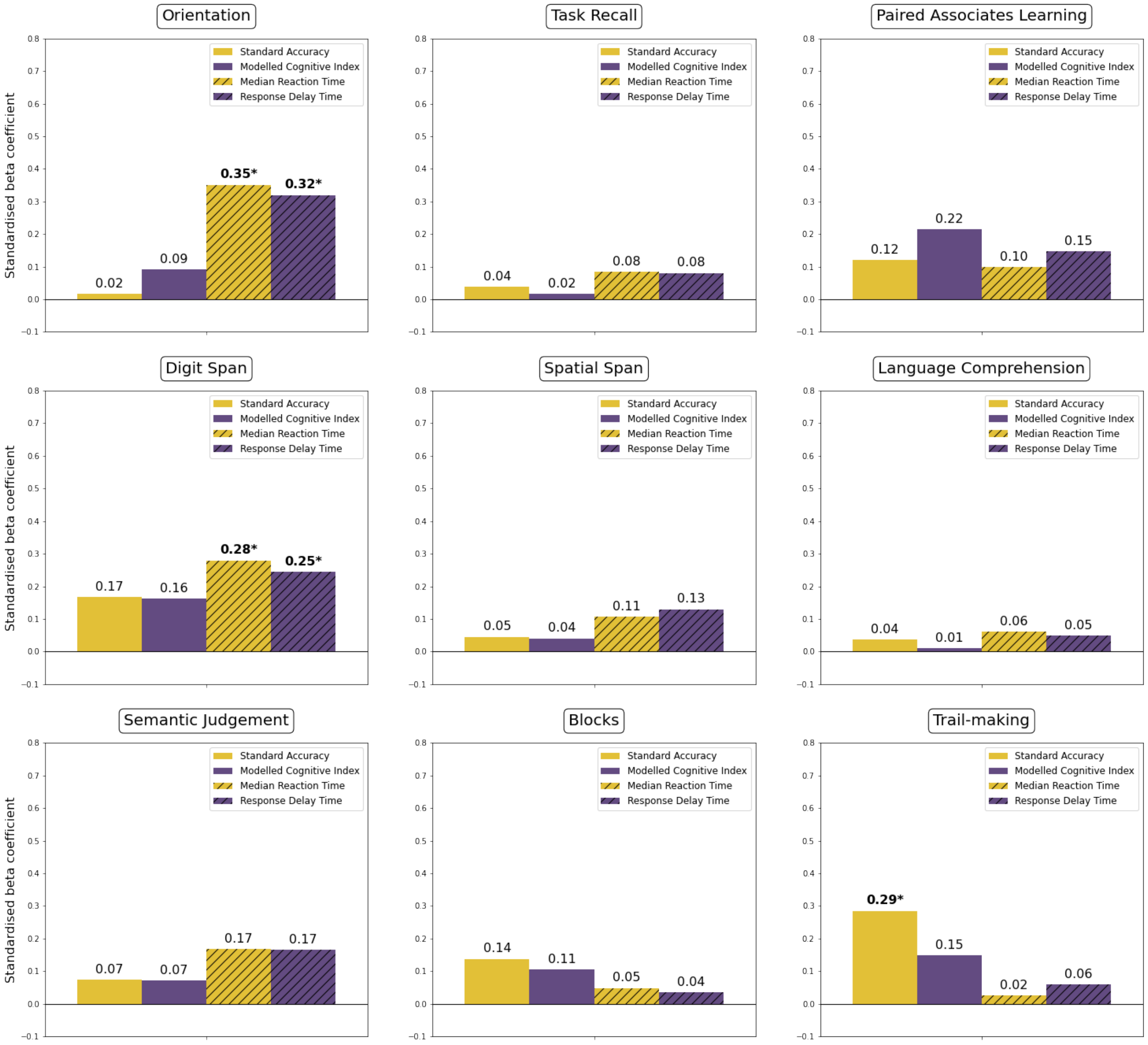

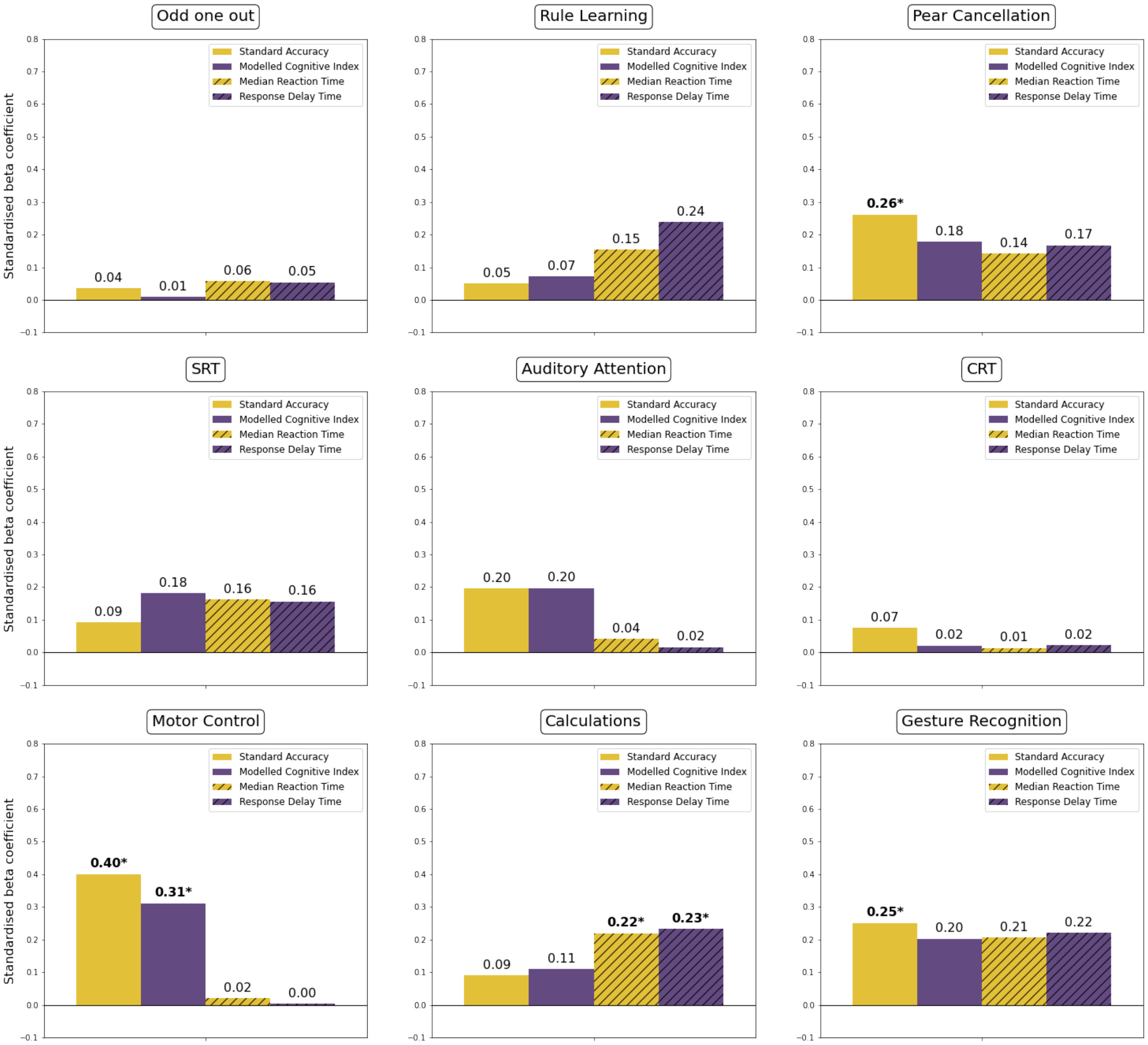
Odd one out
Standard Accuracy
Modelled Cognitive Index
Median Reaction Time
Response Delay Time
Standardised beta coefficient
0.04
0.01
0.06
0.05
Rule Learning
0.05
0.07
0.15
0.24
Pear Cancellation
0.26*
0.18
0.14
0.17
SRT
0.09
0.18
0.16
0.16
Auditory Attention
0.20
0.20
0.04
0.02
CRT
0.07
0.02
0.01
0.02
Motor Control
0.40*
0.31*
0.02
0.00
Calculations
0.09
0.11
0.22*
0.23*
Gesture Recognition
0.25*
0.20
0.21
0.22

**Supplementary Figure 5** – Global cognitive performance estimated via PCA analysis is related to functional outcomes post-stroke (Instrumental Activities of Daily Living-IADL). **Top panel:** Correlation between global standard accuracy metrics ($G_{standard\ accuracy}$) and functional abilities across the acute, sub-acute and chronic phases of the stroke. **Bottom panel:** Correlation between the global modelled Cognitive Index ($G_{cognitive\ Index}$) and IADL scores across the acute, sub-acute and chronic phases of the stroke. P-values are uncorrected.

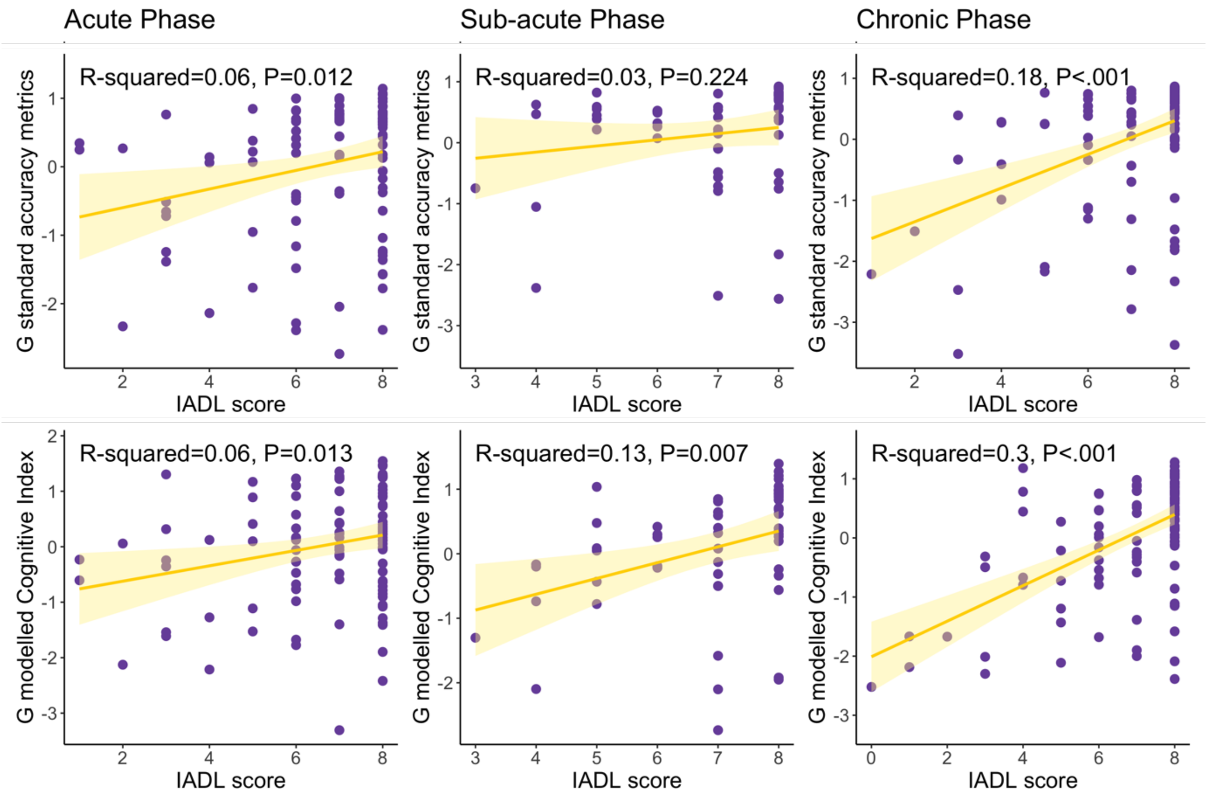

**Supplementary Figure 6.** Lesion overlap map in stroke survivors. Panel A indicates the overlap of the stroke lesion volume, while Panel B indicates the overlap for white matter hyperintensities. A higher overlap of lesion volume across patients is represented in green.

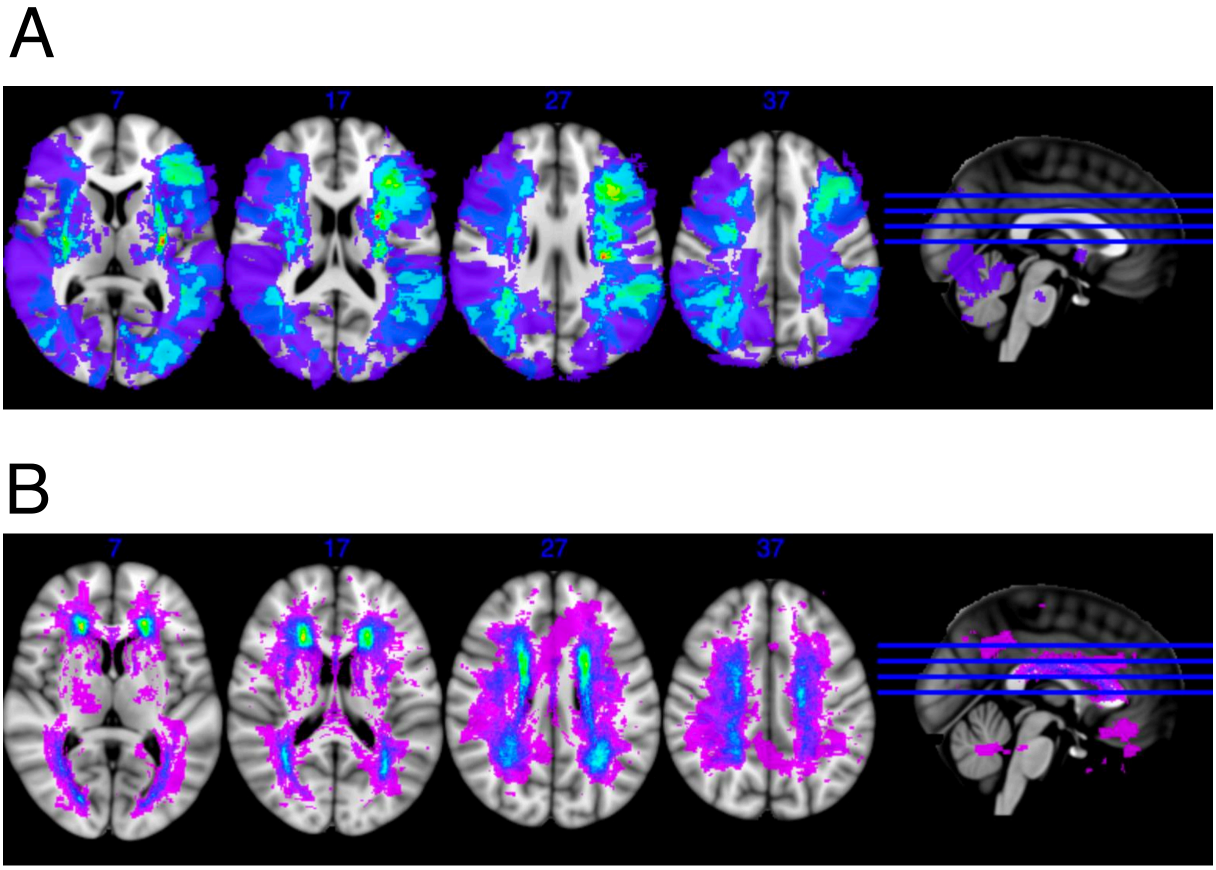
